# Anti-CAR Immunity Drives Acquired Therapeutic Resistance to GD2-CAR T Cell Therapy in Diffuse Midline Glioma

**DOI:** 10.64898/2026.06.25.26356492

**Authors:** Yiyun Chen, Kevin Reynolds, Maximilian R. A. Koch, Camille F. Petrakian, Zinaida Good, Sean Yamada-Hunter, Elena Sotillo, Kun-Wei Song, Jasia Mahdi, Robbie Majzner, Moksha H. Desai, Ying-Wen Huang, Hossein Daghagh, Zachary J. Ehlinger, Nadya Iswari, Chiara Sabatti, Christina Baggott, Skyler P. Rietberg, Kelvin C. Mo, Kristin C.Y. Tsui, Mark P. Hamilton, Emily Egeler, Jennifer Moon, Courtney Erickson, Ashley Jacobs, Allison K. Duh, Barbara Beebe, Casey Carr, Michelle Fujimoto, Michael Kunicki, Alexandria S. Lim, Amy Li, Annie K. Brown, Adam Kuo, Aanchal Kaur, Smriti R. Soundaranayagi, Snehit Prabhu, Gerald Grant, Laura M. Prolo, Cynthia Campen, Sonia Partap, Kara L. Davis, Steven A. Feldman, Ramya Tunuguntla, Jennifer R. Cochran, Bita Sahaf, Sabine Heitzeneder, Michelle Monje, Sneha Ramakrishna, Crystal Mackall

## Abstract

GD2-CAR T cell therapy has demonstrated clinical benefit in patients with H3K27M^+^ diffuse midline glioma (DMG), but the durability of response has been limited in many patients^1,2^. To identify mechanisms of therapeutic resistance, we conducted longitudinal single-cell RNA and TCR sequencing of cerebrospinal fluid (CSF) lymphocytes from DMG patients receiving intravenous followed by sequential intracerebral GD2-CAR therapy, with lymphodepleting chemotherapy administered once prior to the start of CAR T cell therapy (NCT04196413). CSF GD2-CAR T cells manifested limited persistence and clonal expansion, while non-engineered CSF lymphocytes underwent significant clonal expansion and repertoire stabilization, ultimately dominating the CSF immune compartment. Concurrently, peripheral blood CD4^+^ and CD8^+^ T cells manifested anti-CAR immune reactivity targeting epitopes enriched within murine-derived and engineered junctional regions of the CAR construct. This was associated with appearance of circulating Human Anti-CAR Antibodies (HACAs) that bound cells expressing the GD2-CAR, as well as clonal expansion of CSF B cells which produced HACA which impeded the cytotoxic activity of GD2-CAR T cells. In several cases, appearance of circulating HACA temporally correlated with disease progression and across the patient population, and levels of circulating HACA inversely correlated with circulating CAR T cell persistence. These findings reveal robust induction of systemic and CNS adaptive T cell and B cell responses to GD2-CAR T cells following intravenous then sequential intracerebroventricular GD2-CAR therapy and provide strong evidence that anti-CAR immunity is a significant contributor to therapeutic resistance in this setting.

## Introduction

Chimeric antigen receptor (CAR) T cells have revolutionized outcomes for patients with treatment refractory B cell and plasma cell malignancies, inducing durable remissions tantamount to cure in a sizable fraction of patients^3–10^. Recent studies have also reported promising outcomes following CAR T cell therapies for solid tumors^11–13^ and brain tumors^1,2,14–16^, however response rates in these settings remain substantially lower than those observed in B cell and plasma cell malignancies, with very few complete and durable responses reported. These emerging data highlight a critical need to define mechanisms of resistance to CAR T cell therapy for solid tumors and brain tumors in order to guide improved approaches for these patients.

We previously reported clinical results from Arm A of a Phase I trial of intravenous and intracranial GD2-CAR T cell therapy for H3K27M+ diffuse midline gliomas (NCT04196413)^1,2^, wherein 9 of 11 patients treated experienced clinical benefit, four demonstrated major tumor regressions and one patient experienced an ongoing complete response associated with reversal of neurologic deficits, and still durable at the time of writing, now over 4 years since beginning therapy. However, several patients exhibited therapeutic responses for periods of time but then progressed despite continued treatment, suggesting the development of therapeutic resistance. Comprehensive analysis of sequential correlative samples from these patients presented here reveals robust induction of cell-mediated and humoral anti-GD2-CAR immunity, both in the CSF and periphery across the population, which coincides with disease progression in several patients. These data identify immune rejection as an important mechanism of treatment resistance to CAR T cell therapy and demonstrate that CNS immune privilege does not preclude immune rejection of CNS-directed autologous cell therapeutics.

## Results

### GD2-CAR T cells show limited expansion and persistence within the CSF whereas non-CAR T cells and B cells manifest long-term CSF expansion

Thirteen patients with H3K27M^+^ DMG were enrolled on Arm A of NCT04196413; 11 received protocol-directed therapy on study (**Extended Data Fig. 1a**), one received protocol-directed therapy on a compassionate exemption, and one was removed from study prior to receiving therapy due to rapid tumor progression. All treated patients received one cycle of lymphodepletion (LD) (cyclophosphamide 1500mg/m^2^ and fludarabine 75 mg/m^2^) prior to one intravenous (IV) infusion of GD2-CAR T cells. Ten patients who demonstrated benefit based upon magnetic resonance imaging (MRI) and/or neurologic assessment received sequential intracerebroventricular (ICV) infusions without additional LD approximately every 4 weeks until progression or data cut-off on 2/26/24 (cycle number range 1-17, median=5 for 11 treated patients). Based upon volumetric tumor measurements from serial MRIs and neurological assessments, we classified patients as having experienced a sustained response (>12 months), intermediate response (5-12 months) or no sustained response (<5 months) (**Fig. 1a, Extended Data Fig. 1b**, **Supplementary Table 1**). A comprehensive longitudinal single-cell atlas comprising 1,412,518 cells from 181 samples was generated from these patients which comprised 20 CD4/CD8-enriched apheresis samples obtained before (n=12) or during (n=8) therapy, 42 GD2-CAR products sorted into CAR⁺ and CAR⁻ fractions, 113 CSF samples from baseline (n=10, prior to LD and CAR T infusion), predefined and toxicity-mandated post-treatment timepoints (n=103), and 6 samples obtained from an intratumoral cyst (**Supplementary Table 2**). Integrating unsupervised clustering, marker gene identification and reference-guided automatic annotation^17,18^ (see Methods and **Supplementary Table 3**), we catalogued dynamics of lymphoid, myeloid, and tumor cells in the CSF (**Fig. 1b-d**, **Extended Data Fig. 1c-d** and **2a-f**) to identify molecular and cellular mechanisms underlying therapeutic response and resistance. Here we report a detailed analysis of lymphoid cell dynamics while tumor and myeloid population dynamics are described in a companion study (Geraghty, Chen *et al.*, in preparation).

**Fig. 1:**
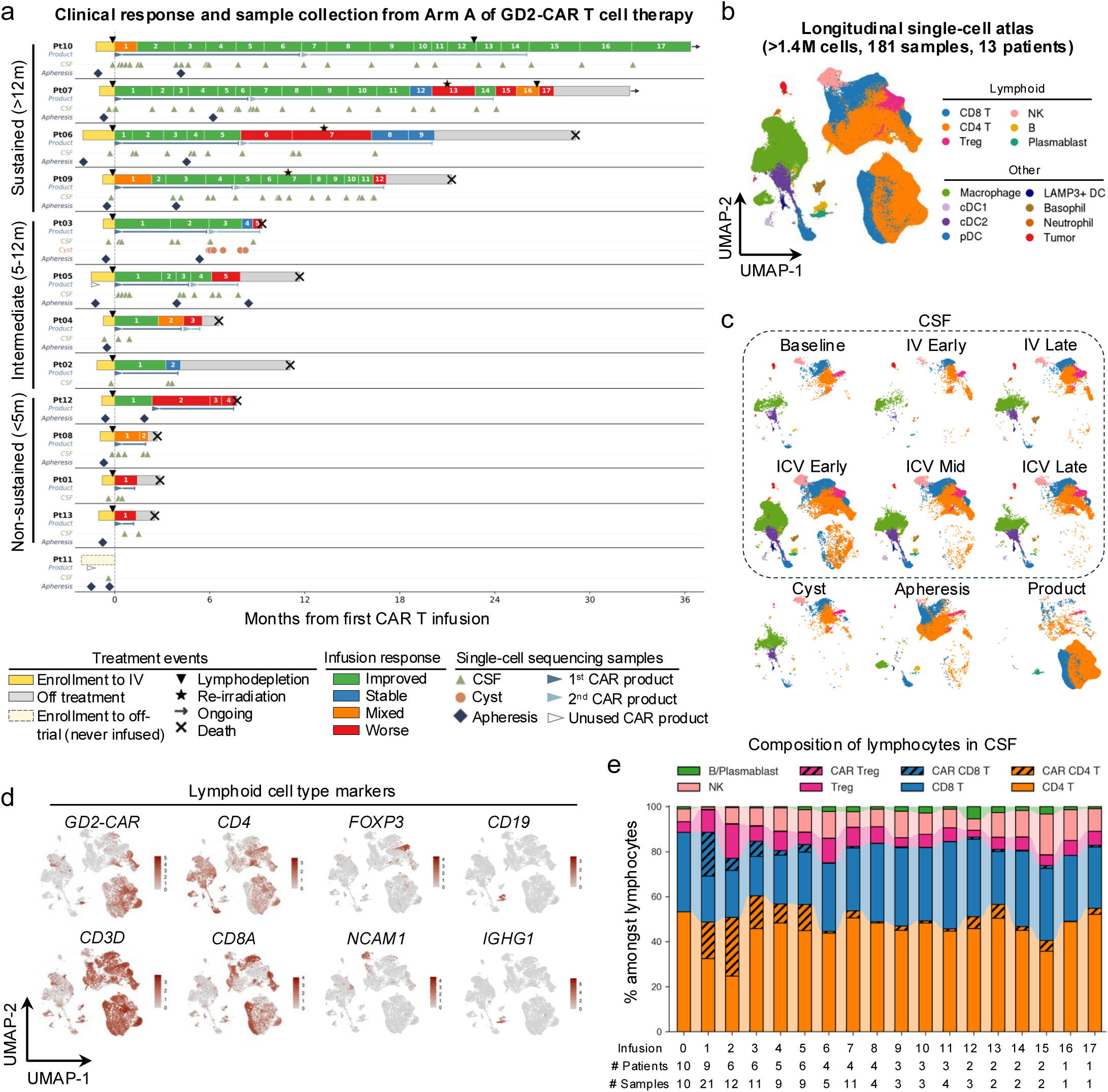
Longitudinal single-cell atlas reveals lymphocytes dynamics in DMG patients during GD2-CAR T cell therapy. **a**, Schematic overview of sample collection for the longitudinal single-cell atlas for 12 GD2-CAR treated H3K27M+ DMG patients enrolled on Arm A of NCT04196413. Eleven were treated on study, while Pt 02 was treated on a compassionate exemption according to the same treatment regimen. Patients are grouped by sustainability of response to the therapy. Product tracks mark the infusion cycles using lots from distinct CAR products. Time relative to the first infusion is marked for CSF, cyst and CD4/CD8-enriched apheresis samples. **b-c**, UMAP visualization of the longitudinal single-cell atlas clustered using the Louvain algorithm as detailed in Methods and colored by annotated cell types in its entirety (b) and across sample types and timepoints relative to GD2-CAR T cell therapy (c). IV-early=D1-14 and IV-late=D15+ after IV infusion. ICV-early, ICV-mid and ICV-late refer to D1-5, D6-14 and D15+ after ICV infusions, respectively. **d**, UMAP visualization of representative lymphocyte cell type markers and the GD2-CAR transgene expression. **e**, Dynamics of indicated cell type fractions among all lymphocytes in the CSF at baseline and at each infusion cycle following the GD2-CAR T cell therapy.

CAR T cells were identified by aligning scRNA-seq reads to a human reference genome augmented with GD2-CAR-specific sequences (**Fig. 1d**), yielding high sensitivity for CAR detection since 86.5% of T cells in CAR⁺ sorted product samples expressed detectable CAR transcripts. Interestingly, 10% of CAR^-^ sorted products revealed CAR mRNA, likely reflecting CAR internalization as previously reported^19,20^ (**Extended Data Fig. 2a**). No CAR^+^ cells were identified in pre-treatment apheresis samples or baseline CSF (**Extended Data Fig. 2a-d**), while on-treatment apheresis samples demonstrated 2% CAR⁺ T cells (originating exclusively from patients Pt05, Pt07, and Pt09; **Extended Data Fig. 2b**). Apheresis and CAR T cell products demonstrated a higher proportion of CD4⁺ relative to CD8⁺ T cells regardless of CAR expression and showed minimal representation of Tregs (**Extended Data Fig. 2b** and **2e**). Following CAR T cell infusion, we observed significant T cell expansion within the CSF, however most cells did not express CAR mRNA (**Fig. 1e**, **Extended Data Fig. 2a, 2c-d and 3a-b**). Furthermore, despite sequential GD2-CAR infusions, we observed lower frequencies of CAR^+^ cells with later cycles compared to earlier cycles (Wilcoxon rank-sum test P=2×10^-6^ between CSF samples from infusions 1-5 versus 6-17) (**Extended Data Fig. 3c-d**). Indeed, following the 6^th^ infusion, levels of CAR^+^ T cells were typically less than 10%, and never higher than 28% of all CSF T cells.

**Fig. 2:**
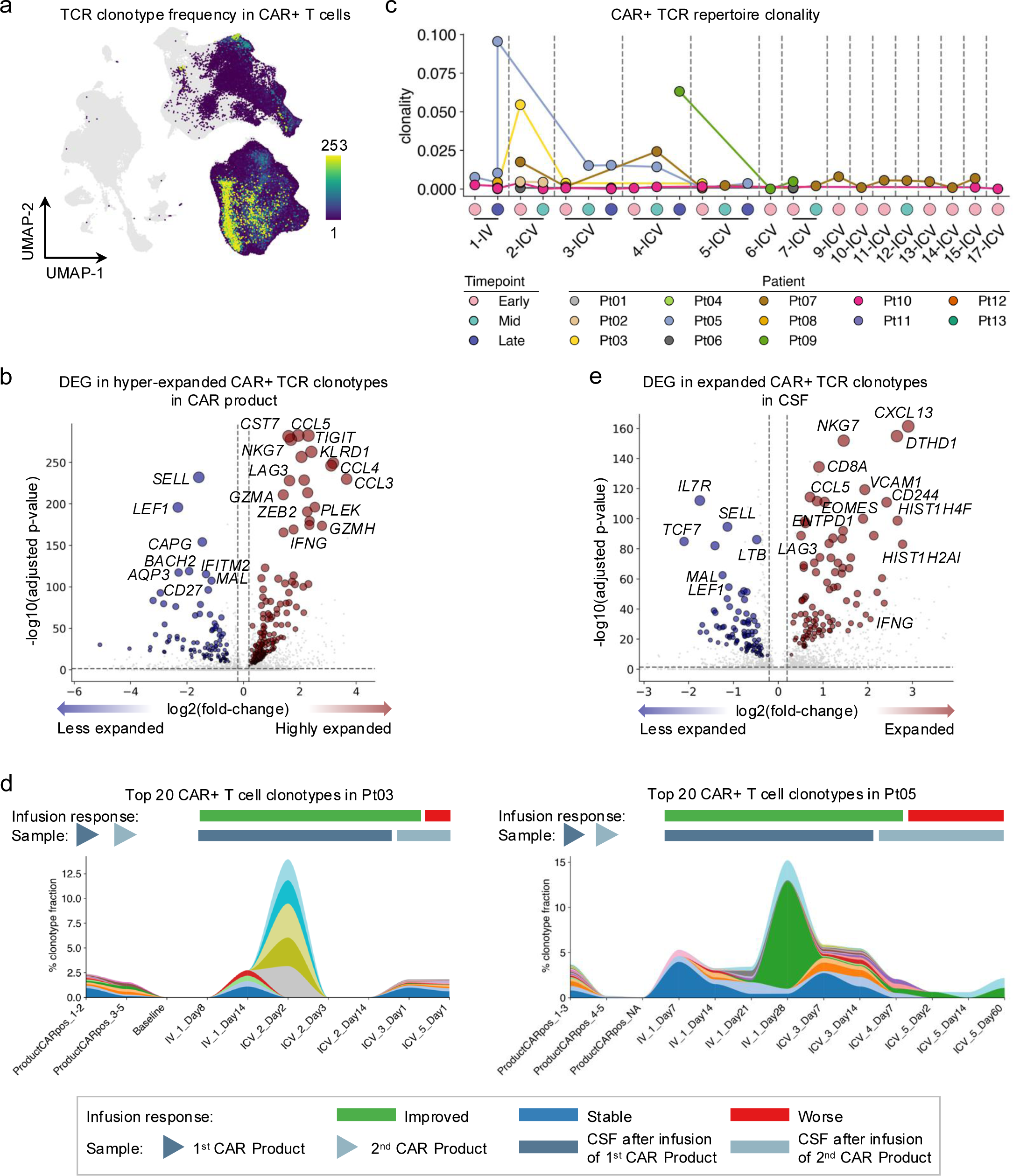
GD2-CAR T cells showed limited expansion and persistence *in vivo*. **a**, TCR clonotype sizes in CAR^+^ T cells. Only CAR^+^ T cells with defined TCR clonotypes were colored, while all other cells in the sample are shown in a grey background. **b**, Volcano plot highlighting top differential expressed genes (DEG) in CAR^+^ CD8^+^ T cells with hyper-expanded (>50 cells per clonotype) compared to less expanded clonotypes (>50 cells per clonotype) clonotypes in the CAR product. X-axis represents the log2(fold-change) of average gene expression level in hyper-expanded versus less expanded clonotypes, and Y-axis represents the-log10(adjusted P-values) by Wilcoxon rank-sum test. **c**, Clonality index of CAR^+^ T cells in CSF samples with at least 100 CAR+ T cells with available TCR information. Each sample is represented as a scatter and colored according to patient ID. The vertical lines mark different infusion cycles. IV-early=D1-14 and IV-late=D15+ after IV infusion. ICV-early, ICV-mid and ICV-late refer to D1-5, D6-14 and D15+ after ICV infusions, respectively. **d**, Clonotype fractions of top 20 CAR+ T cell clonotypes in Pt03 (left) and Pt05 (right). Each color represents one TCR clonotype. The infusion response and corresponding CAR product for each infusion are annotated on top. **e**, Volcano plot highlighting top DEG in CAR+ T cells with expanded (≥3 cells per clonotype) compared to non-expanded clonotypes (<3 cells per clonotype) in the CSF. X-axis represents the log2(fold-change) of average gene expression level in expanded versus non-expanded clonotypes, and Y-axis represents the-log10(adjusted P-values) by Wilcoxon rank-sum test.

In contrast to the low number and frequency of CSF CAR^+^ T cells, substantial frequencies and absolute numbers of CSF CAR^-^ T cells were present throughout the study (**Fig. 1e** and **Extended Data Fig. 2c-d** and **3a-d**). CSF CAR⁻ Tregs, which were rare in apheresis and CAR T cell products, also increased markedly following IV infusion, accounting for up to 40% of total CSF cells (and up to 60% of CD4⁺ T cells), and remained persistently elevated, comprising approximately 10% of total CSF cells (and 20% of CD4⁺ T cells) at later timepoints (**Extended Data Fig. 2d-e**). We also noted small but significant expansions of CSF B cells and plasmablasts at later timepoints (**Fig. 1e, Extended Data Fig. 2f and 3c-d**), whereas CSF NK cell frequencies remained relatively constant throughout treatment. Together, the data demonstrate poor persistence of GD2-CAR T cells within the CSF following one IV and sequential ICV GD2-CAR T cell infusions in contrast to persistent, sizable accumulation of CSF CAR^-^ T cells and episodic late infiltration of CSF B cells and plasmablasts.

### Limited clonal expansion and persistence of CSF CAR⁺ T cells

Single-cell TCR sequencing yielded over 740,000 unique TCR clonotypes across CD4/CD8-enriched apheresis, CAR product and *in vivo* (CSF and cyst) samples. Most clonotypes were only detected in a single sample type, as expected (**Extended Data Fig. 4a-c** and **Supplementary Table 4**). However, 3,389 and 3,430 clonotypes were shared between apheresis and CAR⁺ or CAR⁻ T cells in CAR products, respectively (**Extended Data Fig. 4d**). TCR clonotypes reliably linked each CAR product to its source apheresis, even in patients for whom multiple CAR T cell products were manufactured from different aphereses (**Extended Data Fig. 4e** and **Supplementary Tables 1 and 2**). Overall, quantitative assessment of repertoire uniformity revealed low clonality in most apheresis and CAR product samples (**Extended Data Fig. 4f-g**), although a small subset of CAR^+^ T cells within manufactured products and CSF samples showed clonal expansion (**Fig. 2a** and **Extended Data Fig. 5a**). Dominant CAR⁺CD8⁺ clonotypes (with >50 cells) in products displayed transcriptional signatures associated with effector and exhaustion states, while less expanded clonotypes were enriched for naïve T cell-associated markers (**Fig. 2b**).

Given the low frequency of CSF CAR⁺ T cells, most notable with later infusion cycles (**Fig. 1d**), we sought to determine whether CAR⁺ T cells established durable clonal persistence within the CSF by quantifying TCR clonality among CAR⁺ T cells in CSF samples containing at least 100 CAR⁺ T cells. CAR⁺ T cells exhibited transient increases in clonality in a small number of CSF samples collected during cycle 1-4, however overall clonality remained low (**Fig. 2c** and **Extended Data Fig. 5a**) and further decreased after the fifth infusion, despite repeated dosing. Consistent with this pattern, pairwise TCR repertoire similarity comparison between samples from the same patient using the Morisita-Horn index revealed extremely low overlap of CAR⁺ T cell repertoires across timepoints (**Extended Data Fig. 5b**). Visualization of the most expanded CAR⁺ clonotypes in individual patients confirmed a pattern of episodic, short-lived clonal appearances (**Fig. 2d** and **Extended Data Fig. 5c**). The failure of CSF CAR⁺ T cells to establish durable clonal persistence in CSF is consistent with a model wherein sequential ICV CAR T cell infusions did not induce sustained *in vivo* engraftment but rather continuously reseeded the CSF with new CAR⁺ clonotypes with limited proliferative or survival capacity.

### Significant clonal expansion and durable persistence of CSF CAR⁻ T cell clonotypes

Unlike CAR^+^ T cells, TCR clonotype analysis of CAR^-^ T cells in manufactured cell products and *in vivo* samples revealed extensive clonal enrichment (**Fig. 3a** and **Extended Data Fig. 6a**). Expanded clonotypes spanned multiple T cell states but were most prominent within CD8⁺ T cells and demonstrated progressive clonal amplification over time, as illustrated by marked increase in clonality scores across successive infusions in patients Pt03, Pt05, Pt06, and Pt09 (**Fig. 3b, Extended Data Fig. 6b**). This was accompanied by progressive temporal stability of the CAR⁻ TCR repertoire, with early post-infusion samples showed limited similarity between timepoints, whereas later infusions showed high intra-patient repertoire similarity, as measured by the Morisita-Horn index (**Fig. 3c**). Importantly, CSF TCR repertoires at later infusions were vastly different from the aphereses and CAR products, indicating that these T cells were likely recruited endogenously. Visualization of the top expanded CAR⁻ TCR clonotypes further supported this pattern (**Fig. 3d**), with dominant CAR⁻ clonotypes emerging over later infusion cycles and persisting across multiple subsequent timepoints in patients Pt03, Pt05, Pt06 and Pt09. This pattern was not universal across the cohort (**Fig. 3c and 3d**), as Pt07 exhibited prominent clonal CAR⁻ clonal expansion early during therapy, followed by a loss of dominant clonotypes and reversion to a highly polyclonal repertoire at later timepoints. Similarly, Pt10 showed transient clonal CAR⁻ T cell amplification during the first two infusion cycles, which was not sustained with subsequent dosing. Of note, Pt07 and Pt10 were sustained responders, who remained on study longer than any other in this Arm, and Pt10 continues to manifest an ongoing durable complete response more than four years since beginning therapy. Of interest, a small number of highly expanded Treg clonotypes were also identified in patients Pt05, Pt06, Pt07, Pt09, and Pt10 (**Extended Data Fig. 6b-c**), and these displayed elevated expression of chemokines and immune checkpoint molecules (**Extended Data Fig. 6d**). Together, these analyses demonstrate infiltration and sustained clonal expansion of CAR^-^ T cells in the CSF that stands in stark contrast to the transient and unstable dynamics of CAR⁺ T cells. Interestingly, the marked clonal expansion and progressive repertoire stabilization of CAR⁻ T cells was very pronounced in several patients associated with late clinical progression, while the two patients with the most sustained clinical responses, demonstrated more transient clonal expansion of CAR⁻ T cells, raising the prospect that clonal expansion of CAR⁻ T cells could have contributed to therapeutic resistance.

**Fig. 3:**
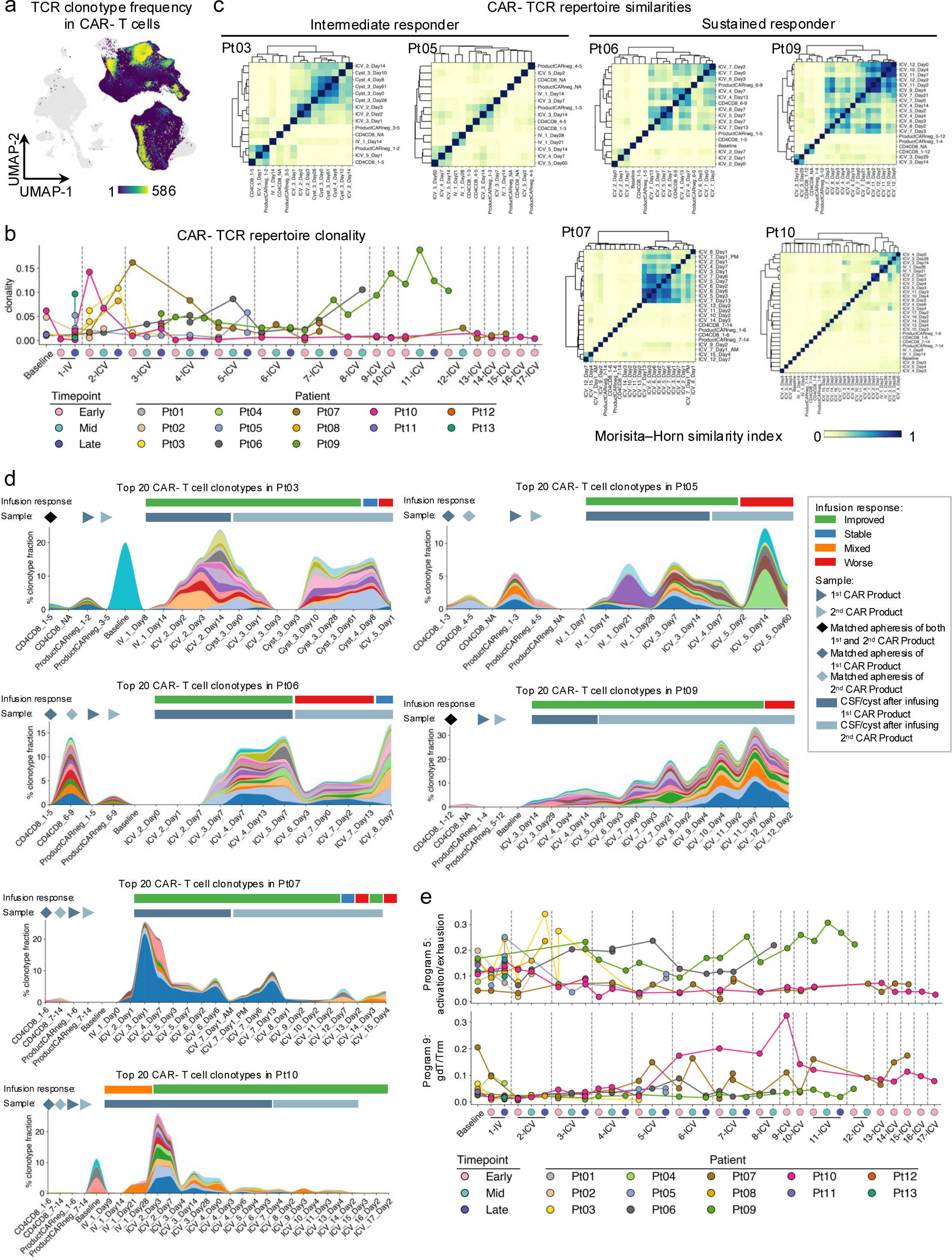
Expansion and repertoire stabilization of CAR-negative T cells *in vivo*. **a**, TCR clonotype sizes in CAR^-^ T cells. Only CAR^-^ T cells with defined TCR clonotypes are colored, while all other cells in the sample are in the grey background. **b**, Clonality index of CAR^-^ T cells in CSF samples with at least 100 CAR^-^ T cells. Each sample is represented as a scatter and colored according to patient ID. The vertical lines mark different infusion cycles. IV-early=D1-14 and IV-late=D15+ after IV infusion. ICV-early, ICV-mid and ICV-late refer to D1-5, D6-14 and D15+ after ICV infusions, respectively. **c**, Pairwise TCR repertoire similarity between samples from the same patient by the Morisita-Horn index. **d**, Clonotype fractions of top 20 CAR^-^ T cell clonotypes in representative patients. Each color represents one TCR clonotype. The infusion response and corresponding apheresis and CAR product for each infusion are annotated on top. **e**, Dynamics of cNMF program 5 (top) and program 9 (bottom) of T cells in across timepoints and patients. Each point represented one CSF sample and was colored by patient ID.

### Transcriptional programs reveal divergent endogenous T cell trajectories following repeated CAR T cell infusions

To define the transcriptional programs associated with the T cell dynamics observed, we performed consensus non-negative matrix factorization (cNMF) across all T cells in apheresis, CSF and cyst, identifying 18 reproducible gene expression programs (**Supplementary Table 5**). A cytotoxic/exhaustion signature (Program 5, marked by *GZMK*, *LAG3* and *EOMES*) aligned with regions populated by highly expanded CAR⁻ clonotypes and accordingly, Pt03, Pt05, Pt06 and Pt09 exhibited progressive enrichment of Program 5 at later infusion cycles (**Fig. 3e and Extended Data Fig. 7a-c**), paralleling increasing TCR clonality and repertoire convergence. In contrast, gamma-delta T cell and tissue-resident memory signatures (gdT/Trm) (Program 9, marked by *ZNF683*, *KLRC2* and *TRDV1*) were enriched in T cells from Pt07 and Pt10, who did not demonstrate progressive CAR⁻ clonal amplification and experienced prolonged disease control. Overall, the average Program 9 score in T cells was significantly higher in patients who showed sustained response to GD2-CAR T cell therapy (**Extended Data Fig. 7d**). Interestingly, ZNF683, a key regulator of Trm and gdT cells^21^, and has been associated with favorable responses to anti-PD1 immunotherapy^22^. We also observed clustering related to temporal dynamics, with enrichment of an NK-like exhaustion signature^23^ (Program 11, marked by *KLRC1*, *KLRD1* and *HAVCR2*) at post-IV timepoints and enrichment of an IFNγ response signature (Program 8, marked by *ISG15*, *IFIT1* and *GBP1*) at ICV-early timepoints (**Extended Data Fig. 7b**). Together, these analyses reveal divergent trajectories of the CAR⁻ T cell compartment among patients: one characterized by tissue-resident or innate-like transcriptional reprogramming without dominant clonal expansion that associated with prolonged disease control and a second characterized by clonal amplification and cytotoxic/exhaustion-associated differentiation.

### Adaptive cellular immunity targets the GD2-CAR transgene

Given the CSF infiltration of clonally expanded CAR^-^ T cells following repetitive CAR T cell infusions and the coinciding loss of CSF CAR^+^ T cells, we hypothesized that GD2-CAR T cell therapy may have triggered antigen-specific immune responses promoting immune-mediated rejection of CAR T cells. To detect cellular immunity against GD2-CAR, we quantified antigen-specific T cell responses by flow cytometry from peripheral blood mononuclear cells (PBMCs) from 5 patients in response to a CAR-spanning 18-mer overlapping peptide library (**Extended Data Fig. 8a-b** and **Supplementary Tables 6-8**; see also Methods). The peptide library covered the entire open-reading frame of the GD2-CAR transgene, including the iCas9-FKBP safety switch, and was organized into 16 pools according to a three-dimensional matrix design^24^, where pools 11-16 correspond to a defined region within the CAR construct (**Extended Data Fig. 8c**).

Compared to healthy donor PBMCs, CD4^+^ T cells from all CAR-treated patients tested showed significant activation-induced marker upregulation and IFN-γ and TNFα production upon stimulation with peptide pools (**Fig. 4a-c** and **Extended Data Fig. 9**). T cells from 4 of 5 patients also demonstrated peptide-induced IL-2 and IL-21 production (**Fig. 4b-c**), while lower levels and frequencies of IL-4 and IL-17A were also detected (**Fig. 4c** and **Extended Data Fig. 8d**). IL-10 was the only measured parameter not produced by CD4^+^ T cells in response to CAR-derived peptide stimulation. CD8^+^ T cells from several patient samples also displayed peptide-induced activation as measured by 4-1BB upregulation and degranulation as measured by CD107a expression (**Fig. 4d-f** and **Extended Data Fig. 8e**). Deconvolution-based epitope mapping of the three-dimensional matrix pool (**Fig. 4g**) identified seven CD4^+^ and five CD8^+^ distinct T cell epitopes, three of which elicited responses in two individuals (**Fig. 4h**). Eight of twelve epitopes eliciting responses localized to the murine-derived 14g2a-based scFv, significantly exceeding the frequency expected by chance (43/150 peptides spanning the scFv; Fisher’s exact test, *P* = 0.0422), and peptides encompassing junction regions within the CAR also manifested increased immunogenicity (5 of 26 peptides) compared with the remainder of the construct (7 of 124 peptides; Fisher’s exact test, *P* = 0.0356). These findings demonstrate that GD2-CAR T cell therapy induces T cell-mediated immune responses comprising both multifunctional CD4^+^ T_H_ cells and cytotoxic CD8^+^ T cells directed against the introduced GD2-CAR and iCas9-FKBP transgene.

**Fig. 4:**
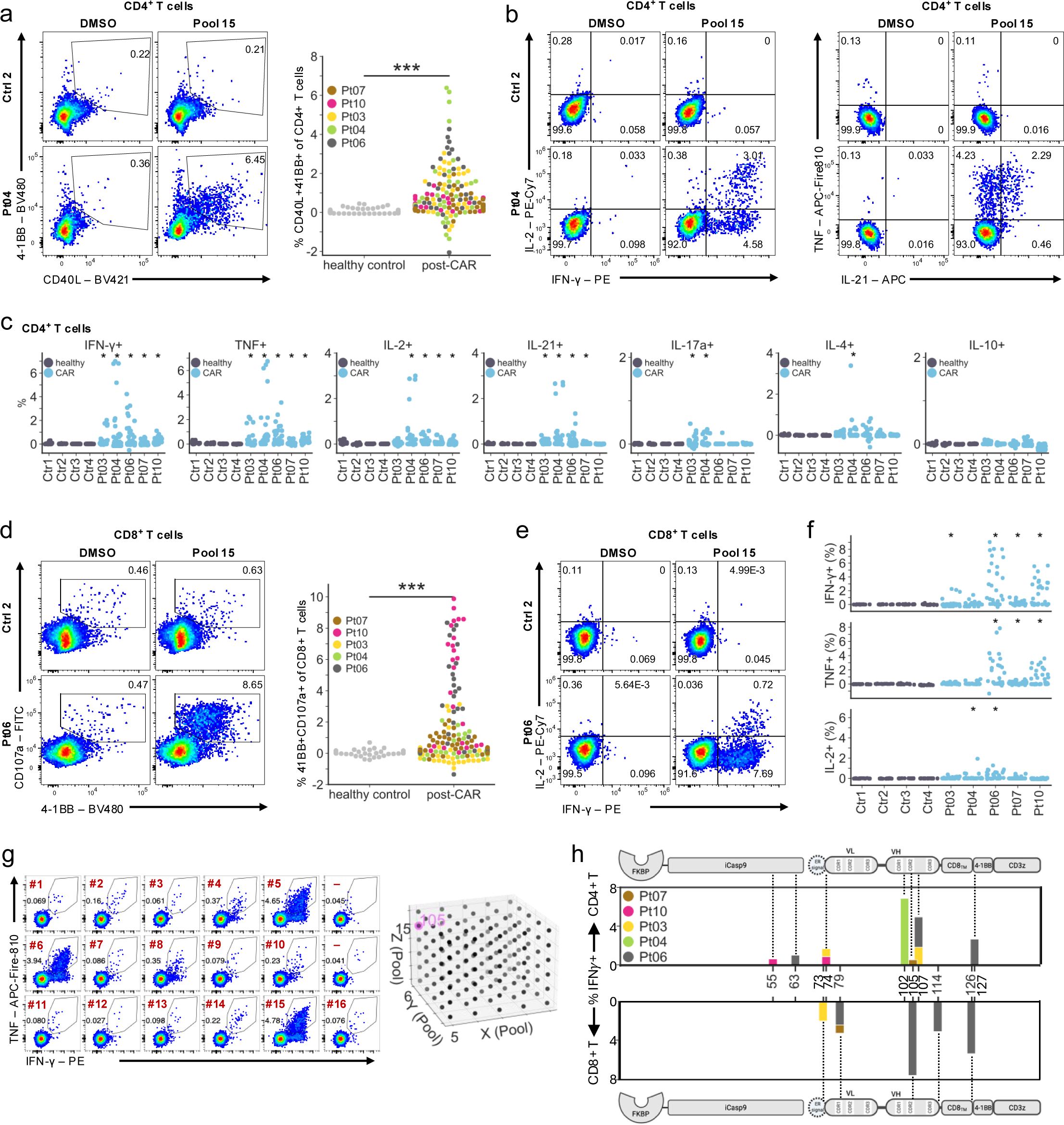
Adaptive cellular immunity targets epitopes on GD2-CAR molecule. **a**, Activation-induced marker expression in CD4⁺ T cells from CAR-treated patients and healthy controls following CAR peptide pool restimulation after antigen-specific in vitro expansion. Left: representative pseudocolor plots show upregulation of CD40L (CD154) and 4-1BB (CD137) in response to pool 15 compared to the DMSO control and a healthy donor. Right: swarm plot summarizes responses across 16 peptide pools for n = 5 patients (two time points each) and n = 4 healthy donors, normalized to DMSO control. \*\*\**p* < 0.001 by linear mixed-effect models. **b**, Representative flow cytometry plots displaying IFN-γ, IL-2, TNFα, and IL-21 secretion of responsive CD4^+^ T cells in the same samples shown as in (a). **c**, Strip plots displaying the DMSO-normalized frequencies (%) of cytokine-positive CD4^+^ T cells following CAR peptide pool restimulation. Each dot represents one peptide pool. Asterisks indicate responses meeting significance criteria based on permutation testing after Bonferroni-correction versus healthy controls and exceeding a twofold increase over DMSO control. **d**, Representative pseudocolor plots (left) and swarm plot (right) showing activation-induced (4-1BB) and degranulation (CD107a) marker expression in CD8⁺ T cells following CAR peptide pool restimulation, analyzed as in (a). \*\*\**p* < 0.001 by linear mixed-effect models. **e**, Representative flow cytometry plots displaying IFN-γ and IL-2 secretion of responsive CD8^+^ T cells in the same samples shown as in (d). **f**, Strip plots displaying the DMSO-normalized frequencies (%) of cytokine-positive CD8^+^ T cells following CAR peptide pool restimulation, analyzed as in (c). Each dot represents one peptide pool. **g**, Representative epitope mapping based on IFN-γ and TNFα production by CD8⁺ T cells from one patient following peptide pool restimulation. Flow cytometry plots show responses to 16 overlapping peptide pools and two DMSO controls (–). Pools 5, 6, and 15 elicit comparable cytokine responses, allowing assignment to peptide 105 based on the three-dimensional matrix design. **h**, Relative positions (Peptide ID) and IFN-γ response strength of all via epitope mapping identified CD4^+^ (top) and CD8^+^ (bottom) T cell epitopes on GD2-CAR and iCas9-FKBP proteins.

### Neutralizing anti-CAR antibodies emerge in the plasma and CSF of patients receiving GD2-CAR therapy and associate with therapeutic resistance

Our single-cell atlas revealed expansion of CSF B cells and plasmablasts in several patients (**Fig. 1e** and **Extended Data Fig. 2f**), and further analysis of the CSF B cell receptor (BCR) repertoire using the TRUST4 algorithm^25^, revealed several highly expanded BCR clonotypes, most notable in Pt06, Pt07 and Pt09 (**Supplementary Table 9**), raising the prospect that induction of humoral anti-CAR immunity within the CNS contributed to treatment resistance. To test this, we correlated treatment response with levels of CSF B cells, plasmablasts and B cell clonality at each treatment cycle in individual patients. As shown in **Fig. 5a**, across all infusion cycles, those associated with worse clinical response showed a trend for temporal increases in CSF B cell fractions (two-sided Wilcoxon test, P = 0.062). We also observed several patients for whom increased CSF B cell or plasmablast frequency and/or CSF B cell clonality immediately preceded or coincided with clinical progression (**Fig. 5b**). Of note, Pt10, who experienced an ongoing complete response, exhibited minimal CSF B cell expansion (**Fig. 5a-b** and **Extended Data Fig. 2f**).

**Fig. 5:**
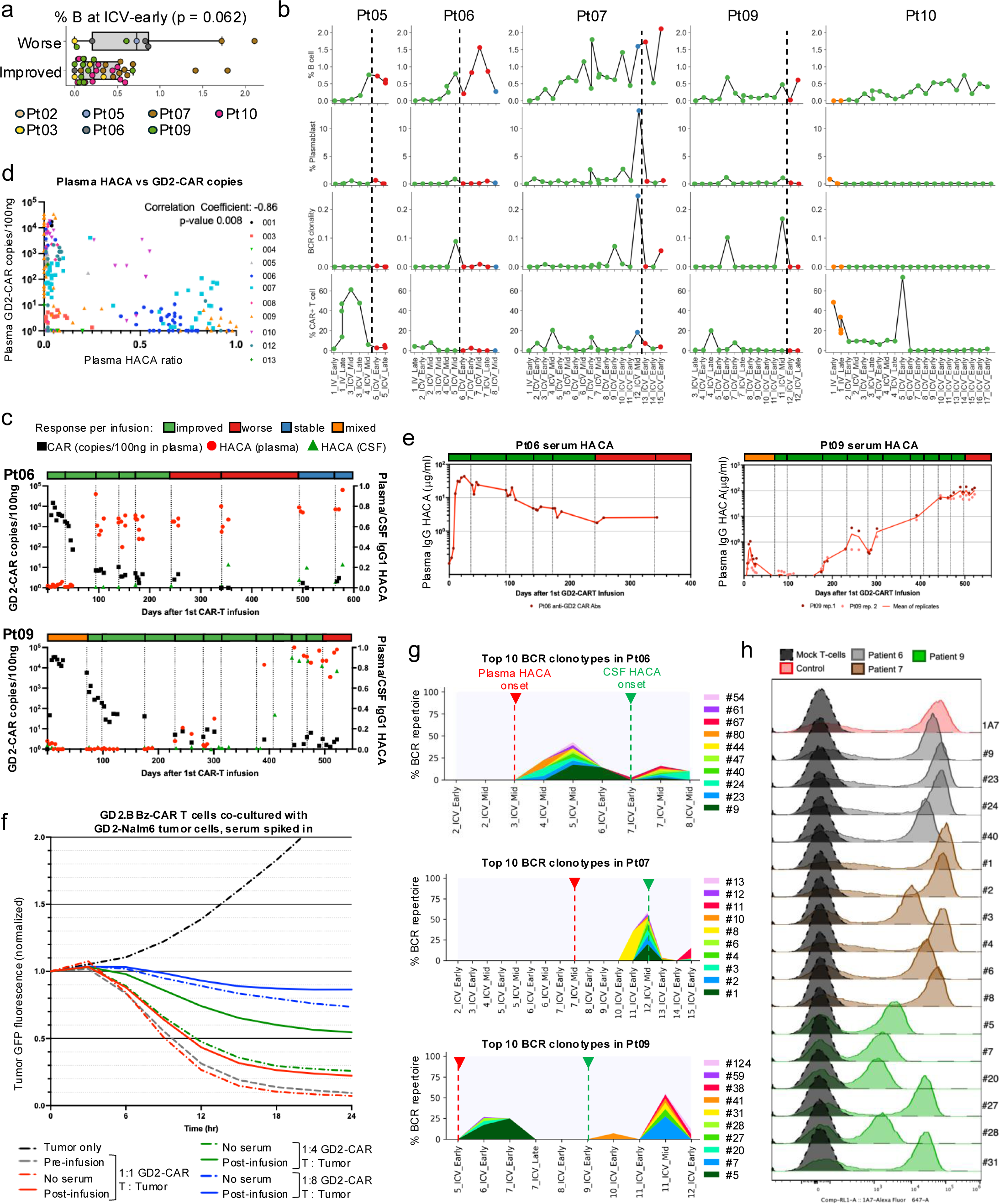
B cells expansion and HACAs oppose CAR T cell persistence and activity. **a**, Percentage of B cells in scRNA-seq data of CSF samples collected at ICV-early timepoint of infusion cycles during which patients show improved versus worse response to GD2-CAR T cell therapy. P-value by Wilcoxon rank-sum test. **b**, From top to bottom panels: percentage of B cells, plasmablast, BCR clonality estimated from TRUST4-reconstructed BCR repertoire, and percentage of GD2-CAR+ T cells in scRNA-seq data of all CSF samples from each patient (columns) temporally ordered and plotted. Point colors represent the response at the end of each infusion cycle. The dotted vertical line marks timepoint of switched response from improved or stable to worse. **c**, Longitudinal profiling of GD2-CAR copies (left axis) and human anti-CAR antibodies (HACAs) ratio (right axis) in plasma and CSF of Pt06 and Pt09 by flow cytometry assay. Response per infusion is indicated on top of each graph. **d**, Pearson’s correlation coefficient between plasma GD2-CAR copies and plasma HACA ratio. all samples are color coded by patient. **e**, Longitudinal quantification of HACA levels in Pt06 and Pt09 plasma measured via ELISA. Plasma HACA concentrations (µg/ml) are plotted on a logarithmic scale against days following the initial GD2 CAR-T infusion. For Pt09, individual replicates (symbols) are shown alongside the mean (solid line). Dotted vertical lines indicate each CAR T cell infusion. **f**, Live-cell imaging analysis evaluating the survival of GFP-expressing GD2-Nalm6 target cells during co-culture with GD2.BBz CAR-T cells. Target cell viability was monitored every 4 hr using an IncuCyte and is plotted as tumor GFP fluorescence normalized to the initial time point (t = 0) over a 24-hour period. The black dashed-dotted line represents the baseline growth of tumor cells cultured alone. Red lines represent 1:1 E:T ratios, green represents 1:4, and blue 1:8. Data represent the mean of n=3 replicates. Dashed, colored lines denote co-cultures spiked with 1:1000 high-HACA Pt09 serum, and solid lines represent cocultures with no added serum. Red lines represent 1:1 E:T ratios, green represents 1:4, and blue 1:8. Data represent the mean of ratios with n=3 replicates. **g**, Percentage of the top 10 expanded BCR clonotypes in CSF across treatment timeline in each patient. The BCR sequences were reconstructed with TRUST4 pipeline. Dotted lines marked the time of onset of HACAs in plasma (red) and CSF (green). **h**, Flow cytometry histograms showing binding of recombinant single patient antibody clone supernatants to mock T cells (black) or human primary GD2-CAR T cells (colored). Anti-GD2 CAR idiotype 1A7 was used as positive control (red). Individual clones are colored by patient.

To identify antibodies reactive toward the GD2-CAR T cells, we incubated GD2-CAR expressing Jurkat cells with plasma and CSF samples collected at multiple timepoints across 11 patients (**Fig. 5c**, **Extended Data Fig. 9a** and **Supplementary Table 10**) then quantified the level of bound IgG using an anti-IgG1 antibody. Results seen following incubation with saturating amounts of a control anti-idiotype antibody targeting the GD2-CAR to provide a positive control. Among 4 sustained responders who received at least 6 GD2-CAR infusions, all manifested high levels (>60% neutralization achieved with control anti-idiotype) of IgG plasma <u>H</u>uman <u>A</u>nti-<u>C</u>AR <u>A</u>ntibodies (HACAs), the onset of which temporally coincided with disease progression in 3 patients (Pt06, Pt07 and Pt09). Of interest, Pt10, who experienced an ongoing complete response, developed moderate levels of plasma HACAs (10-60% of neutralization observed with control anti-idiotype) compared to other patients who ultimately progressed. Amongst 7 patients who received less than 6 infusions and for whom HACA data is available, 4 developed moderate HACA levels (**Extended Data Fig. 10a**). Onset of HACAs occurred following at least 3 treatment cycles in all patients except Pt12, where significant levels were detected following the first treatment cycle. Altogether, plasma HACA levels negatively correlated with GD2-CAR qPCR signal in plasma (**Fig. 5d**, coefficient-0.86, SD 0.32, p-value 0.008) even when accounting for time from initial GD2-CAR T cell infusion, supporting an association between anti-CAR antibody development and diminished circulating CAR T cell persistence.

To more accurately quantify HACAs and further analyze their specificity in these patients, we tested reactivity of serum samples from Pt06 and Pt09 against using a HACA-ELISA assay designed to detect and quantify IgG1 antibody binding to plate-bound recombinant scFv derived from the GD2-CAR. The ELISA detected HACAs at concentrations as high as 100μg/ml with kinetics that mirrored those observed by flow cytometry (**Fig. 5e** and **Supplementary Table 11**). To test if circulating HACAs could interfere with the anti-tumor activity of GD2-CAR T cells, we co-cultured human GD2-CAR T cells and Nalm6-GD2^+^ cells (a leukemia cell line engineered to express GD2) in the presence of Pt09 serum (at 1:1000 dilution) collected at HACA-negative (pre-infusion) versus HACA-peak (day 490 from the first infusion) timepoints. HACA-peak serum significantly diminished the cytotoxic activity of GD2-CAR T cells at every effector to target ratio tested (**Fig. 5f** and **Extended Data Fig. 10b**).

The detection of peripheral T cell and antibody-mediated anti-CAR responses associated with CSF T cell clonal expansion and B cell clonal expansion raised the possibility that anti-CAR immune responses could originate or be sustained by lymphocytes within the CSF. We thus tested the CSF of 8 patients using our cell-based assay to identify IgG1 HACA in the CSF. CSF IgG1 HACAs were detected in Pt09 at similar levels as found in plasma, whereas in Pt06 and Pt07 where CSF IgG1 HACAs levels were lower than those found in the plasma, and CSF IgG1 HACAs were not detected in the other patients (**Fig. 5c** and **Extended Data Fig. 10a**). We concluded that levels of IgG1 HACAs were higher and more prevalent in the plasma than the CSF in most patients, suggesting that IgG1 HACAs likely originated and were sustained by extra-CNS B cells and/or plasma cells.

We sought to further interrogate whether CSF B cells produced anti-CAR antibodies, by focusing on Pt06, Pt07 and Pt09 for whom plasma HACAs levels were high and the relationship between HACAs emergence and loss of GD2-CAR persistence was strikingly notable. We thus reconstructed BCR clonotypes from all CSF scRNA-seq samples and discovered that most clonally expanded CSF B cells encoded immunoglobulin A1 (IgA1), followed by IgA2, IgG1 and IgG2 (**Fig. 5g, Extended Data Fig. 11a** and **Supplementary Table 9**). Single-cell BCR sequencing of 10 samples from selected patients validated the presence of these clonotypes, enrichment of IgA isotype, and a significant increase in BCR clonality over time (**Supplementary Table 12**). To test if these antibodies recognize and bind to GD2-CAR T cells, we recombinantly expressed paired variable and lambda or kappa light chains in a human IgG1 format, validated their expression by anti-human Fc ELISA (**Extended Data Fig. 11b**). All 13 antibodies tested bound to human GD2-CAR T cells by flow cytometry, while none bound to mock engineered T cells (**Fig. 5h**). Collectively, these data confirm that CSF B cells produce HACAs targeting GD2-CAR T cells and this adaptive immune response opposes CAR T persistence and interferes with antitumoral cytotoxic activity. The finding that most of the CSF HACA was an IgA1 and IgA2 isotype potentially explains the low level of CSF HACA identified using our cell based HACA assay, which only detected IgG1 antibody responses.

Given the robust B cell responses against GD2-CAR, we reasoned that these responses were supported by cognate T cell help^26^. Notably, stratification of the samples used to detect anti-CAR CD4^+^ T cell responses revealed that these responses were detectable and even peaked prior to the emergence of HACAs (**Fig. 6a-b**). This pattern was even more extreme in the CD8^+^ T cell subsets (**Fig. 6a** and **6c**). Anti-CAR CD4^+^ and CD8^+^ T cell responses developed in the circulation as early as within two weeks post-infusion and subsequently declined in magnitude over time (**Fig. 6d** and **Supplementary Table 13**). Integration of cellular and humoral immune measurements across patients delineated a consistent temporal sequence of anti-CAR immune activation (**Fig. 6d** and **Extended Data Fig. 11c**), wherein early induction of anti-CAR T cell responses was followed by the emergence of circulating HACAs in the periphery, which subsequently became detectable within the CSF at later timepoints and at lower levels. This stepwise evolution of anti-CAR immunity temporally aligned with reduced CAR T cell persistence, diminished functional activity, and eventual clinical progression. The coordinated temporal pattern of anti-CAR immunity provides a mechanistic framework linking immune recognition of CAR T cells to their progressive loss of persistence and therapeutic efficacy and suggests that mitigation of both cellular and humoral immunogenicity will be necessary to improve the durability of CAR T cell therapy in solid tumors.

**Fig. 6:**
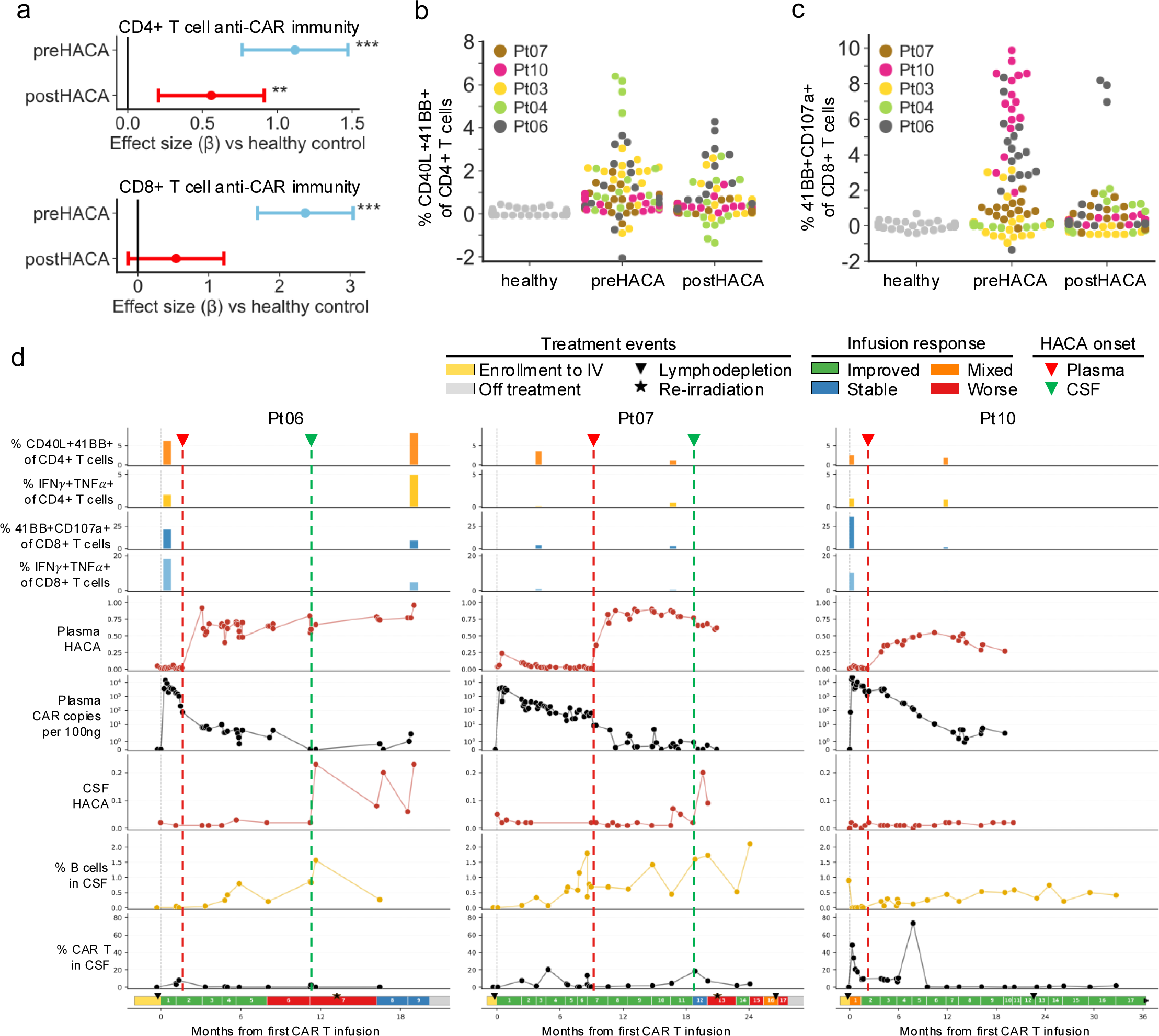
**Anti-CAR cellular immunity emerges prior to humoral immunity** a, Linear mixed-effect model analysis of activation-induced marker expression in CD4⁺ T cells (top) and CD8⁺ T cells (bottom) from CAR-treated patients stratified by timepoint relative to HACA emergence following CAR peptide pool restimulation compared to healthy controls. Swarm plot groups the same datapoints as shown in Fig. 4b and 4c for CD4⁺ T cells and CD8⁺ T cells, respectively. Forrest plot gives the estimated fixed-effect coefficient between CAR-treated groups and healthy controls; points indicate fixed-effect estimates (β), with 95% confidence intervals and associated p-values; \*\**p* < 0.01, \*\*\**p* < 0.001. **b-c**, Anti-CAR CD4⁺ T cells (b) and CD8⁺ T cells (c) response magnitude (aggregated DMSO-normalized response against all 16 pools divided by 3) vs. days post initial CAR infusion. A star indicates a value significant higher (>3SD) than mean of healthy controls. **d**, Longitudinal immune and clinical profiles of three patients with sustained responses to GD2-CAR T cell therapy. The first four tracks capture anti-CAR T cell immune responses measured in peripheral blood. Specifically, the percentages of CD40L+41BB+ and IFNγ+TNF+ cells among CD4+ T cells (tracks 1-2, orange bars) and CD8+ T cells (tracks 3-4, blue bars) quantified at each timepoint by intracellular cytokine stimulation assay. Plasma HACAs (track 5, red) and CSF HACAs (track 7, red) levels as measured by flow cytometry. Plasma CAR T cell copy number per 100ng (track 6, black) as measured by qPCR. Percentages of B cells (track 8, yellow) and CAR+ T cells among total cells in CSF (track 9, black) were derived from scRNA-seq data. The clinical timeline at the bottom of each column indicates the treatment course, with colored segments denoting response per infusion cycle. The dashed vertical red and green lines mark the approximate onset of HACAs in plasma and CSF, respectively.

## Discussion

Anti-drug antibodies are well known mediators of therapeutic resistance^27^. CAR immune cell therapies, the first large scale therapeutic deployment of entirely synthetic proteins have long been recognized to pose a risk for anti-CAR immune responses. While anti-CAR T cell immune responses have been sporadically reported^12,28–30^ convincing evidence has not demonstrated that these contribute, in a significant way, to treatment resistance in the setting of autologous CAR T cell therapies. Here we provide substantial evidence that autologous GD2-CAR T cell therapy, administered via a regimen incorporating one cycle of lymphodepletion, one IV GD2-CAR infusion and sequential ICV infusions, reproducibly induces robust CD4^+^ and CD8^+^ mediated anti-CAR responses and anti-CAR antibodies, both in the CNS and in the periphery (**Fig. 4/5, Extended Data Fig. 8/9**). Anti-CAR T cell responses targeted the murine scFv as well as junctional regions of the CAR receptor and fully human regions within iCas9 (**Fig. 4h**). Our data strongly suggests that these responses contribute to therapeutic resistance based upon: i) limited persistence of CSF GD2-CAR T cells coincident with clonal expansion of non-CAR CSF T cells (**Fig. 1e/3d, Extended Data Fig. 2d**); ii) temporal alignment between appearance of anti-CAR antibodies and disease progression in individual patients (**Fig. 5c, Extended data Fig. 9**); iii) inverse correlations between levels anti-CAR antibodies and GD2-CAR persistence across the population (**Fig. 5d**); iv) inhibition of CAR mediated tumor killing by anti-CAR antibodies (**Fig. 5f**). We anticipate that additional evidence to confirm the role of anti-CAR responses in therapeutic resistance will be gleaned by comparing outcomes from patients treated on Arm A to patients currently being enrolled on arms delivering an intensified lymphodepletion regimen designed to reduce or eliminate anti-CAR immune responses.

Most patients who have received investigational and commercial CAR T cells to date have been treated for refractory B cell and plasma cell malignancies and therefore were highly immunosuppressed at the time CAR T cells were administered. Further, all commercial CAR T cells today target CD19 or BCMA and thus eradicate B and plasma cells respectively, limiting induction of anti-CAR antibody responses^31^. Increasingly however, CAR immune cells that do not deplete immune populations are being tested in patients with brain tumors and solid cancers^11–13,15,16,32–34^, populations with less disease-associated immunosuppression, where the risk of immune rejection may be higher. In support of this concept, a recent trial of CAR T cells targeting Claudin 18.2 in gastric malignancy reported that 95% of CAR T cell recipients demonstrated anti-CAR antibodies^12^. Furthermore, substantial investment is underway to eliminate the use of lymphodepleting chemotherapy prior to CAR T cell infusion with the goal of reducing treatment related morbidity^35^, especially for patients with autoimmune disease and those receiving *in vivo* CAR T cell therapy^36,37^. The results presented here raise the prospect that anti-CAR immunity may emerge as an important cause of treatment resistance in these settings. Immune rejection has limited the success of allogeneic CAR immune cell therapies thus far^38^, leading to the development of numerous technologies to minimize immunogenicity and/or evade the host immune response^39–45^. Our results suggest that incorporating stealth technologies into autologous CAR T cell products could also potentially hold value.

Historically, the CNS has been described as “immune-privileged”^46^, characterized by limited immune cell ingress and egress, leading many to assume that anti-CAR immune responses would be insignificant in the setting of cell therapy for brain tumors. However, recent studies have identified extensive neuroimmune crosstalk, including via neuroimmune hubs within the meninges and choroid plexus, meningeal lymphatics and CNS adjacent tissues such as skull, vertebral bone marrow and deep cervical lymph nodes^47–50^. Significant peripheral:CNS crosstalk has been directly observed in the context of CAR T cell therapy for leukemia, lymphoma and brain tumors, where intravenously administered cells traffic to the CSF and enter the CNS parenchyma^28,33^, and vice versa, where ICV administered CAR T cells traffic into the periphery in animals and patients with brain tumors^2,51^. The findings presented here illuminate remarkably robust induction of T cell-mediated and humoral immune responses within the CNS and periphery in response to a regimen comprising systemic and intracranial administration of autologous CAR T cells, consistent with the evolving paradigm-shift in our understanding of CNS immune privilege. Data demonstrating that a sizable fraction of the anti-CAR antibody producing CSF B cells produce IgA (**Extended Data Fig 11a**) aligns with data demonstrating that meninges are rich in gut educated IgA producing plasma cells, which play a central role in protecting the CNS from pathogens^52^. These results are in support of a model whereby at least a portion of the immune responses induced in this setting originate within the CNS or CNS border tissues.

CAR T cells for solid cancers have shown promise, but high rates of durable and complete remission have not yet been observed^12,13,15,16,32–34^. A holy grail of immunotherapy is induction of endogenous immunity, often described as epitope spreading, wherein long-lasting, individualized immune responses to tumor-specific and tumor-associated antigens mediate durable long-term immunity^53–55^. There is limited evidence thus far that CAR T cell therapies induce significant epitope spreading and no clinical studies have demonstrated a role for this phenomenon in disease control. While the anti-CAR immune responses observed here contributed to immune resistance, the magnitude and duration of these responses suggest that CAR T cells could be exploited to induce high level, durable immunity to neoantigens and tumor-associated antigens, which could improve tumor control. Future studies are needed to determine whether and how to combine CAR T cells with tumor vaccination to improve patient outcomes.

Previous studies focusing on resistance to CAR T cell therapies have emphasized important roles for tumor heterogeneity, T cell exhaustion and dysfunction related to the immunosuppressive tumor microenvironment, and impaired T cell trafficking^56^. Our data demonstrate that resistance to GD2-CAR T cell therapy in diffuse midline gliomas is driven, at least in part, by robust induction of T cell and B cell mediated anti-CAR immunity, highlighting a need for increased evaluation of immune rejection as a mechanism of resistance to CAR immune cell therapies more broadly and demonstrating that CNS immune privilege does not preclude immune rejection of CNS directed autologous cell therapeutics. These results also provide a note of caution for efforts aimed at eliminating lymphodepletion and raise the prospect that stealth engineering could provide important benefits in autologous CAR T cell therapeutics.

### Patient sample processing

Samples were collected from patients with H3K27M-mutant DMG enrolled in the Phase I GD2-CAR T trial (NCT04196413) after informed consent. The patient ID is consistent with Monje *et al.*^2^. Pre-lymphodepletion and post-treatment peripheral blood mononuclear cells (PBMCs) were obtained by leukapheresis and enriched for CD4+ or CD8+ T cells. Cryopreserved GD2-CAR T cell products were thawed then physically sorted into CAR+ and CAR-populations using an anti-idiotype antibody (clone 1A7)^57^ that recognize the 14G2a-derived scFv region of GD2-CAR, resulting in purity of >98% CAR+ or CAR– populations, respectively. CSF was sampled via Ommaya reservoir at protocol-defined intervals (e.g. days 0, 3, 7, 14, 28, etc.) and toxicity-mandated timepoints, and was immediately processed. Patient Pt03 developed a treatment-associated intratumoral cyst during the third and fourth infusion cycles, and 6 cyst fluid samples were obtained at day 0, 3, 10, 28, 61 after the third infusion and at day 8 after the fourth infusion and immediately processed. CSF and cyst fluid cells were pelleted by gentle centrifugation, resuspended in buffer, and live cells counted for downstream assays.

### Single-cell RNA, TCR and BCR sequencing

Single-cell RNA sequencing (scRNA-seq) was performed using 5’ v2 Single Cell Immune Profiling technology (10X Genomics) according to the manufacturer’s protocol. In brief, cells from CD4/CD8-enriched apheresis, CAR product, CSF and cyst fluid samples collected at indicated timepoints before and after CAR T cell administration were counted, resuspended to 700-1,200 cells per µl, and captured using Single Cell Chip A on the 10x Chromium Controller (10X Genomics) to generate gel bead-in emulsions (GEMs). Reverse transcription inside GEMs was performed using a C1000 Touch Thermal Cycler (Bio-Rad). Barcoded complementary DNA (cDNA) was recovered through post-GEM-RT cleanup and PCR amplification. Recovered cDNA was amplified and used to construct 5’ whole-transcriptome and V(D)J T cell receptor libraries. For some samples, BCR V(D)J libraries were also made to identify BCR sequences of CSF B cells. Quality of cDNA and each library was assessed using Agilent 2100 Bioanalyzer. The libraries were indexed using a Chromium i7 Sample Index Kit, pooled and sequenced on NovaSeq 6000 System (Illumina) by Novogene.

### Single-cell data processing, filtering, dimensionality reduction and cell type annotation

Raw sequencing data were processed using the Cell Ranger software version 7.2.0 (10X Genomics). Sequencer’s base call files (BCLs) were demultiplexed into FASTQ files using the cellranger mkfastq pipeline. The GRCh38 5.0.0 human genome reference with GD2-CAR sequence developed previously^1^ was used in the cellranger count pipeline to align the FASTQ files and generate unique molecular identifiers (UMIs) counts. FASTQ reads for scTCR-seq and scBCR-seq were aligned to the vdj-GRCh38 reference version 7.1.0 using the cellranger vdj pipeline.

UMI count matrices were imported into Seurat (v5.3.0) for analysis. Dead cells and cell debris with more than 10% of UMI counts mapping to mitochondrial genes, less than 500 genes or 1,000 UMIs detected were excluded from the analysis. Cell doublets containing more than 5,000 genes or more than 100,000 UMI counts were also excluded. Data were log-normalized using the NormalizeData function. PCA was performed on top 2,000 variable genes except for *TCR* and *BCR* genes to prevent clonotypes from driving the final layout, and the first 30 principal components were then used for UMAP embedding. Cell clusters were generated based on the shared nearest neighbor network built on the first 30 principal components and with Louvain algorithm at the resolution of 0.4. Differential expression analysis of cell clusters was performed using the FindAllMarkers function with Wilcoxon rank-sum test. Cell types were assigned to each cluster based on canonical lineage marker expression and guided by automatic cell type annotations using Azimuth with human PBMC dataset^18^ as reference and SingleR^17^ with Monaco dataset^58^ as reference. The H3K27M mutation was detected using the cellsnp-lite package^59^, and H3K27M-mutant cells were defined as cells with at least 3 UMIs with H3K27M mutation. The cell cycle phase was predicted using the CellCycleScoring function in Seurat. Louvain clustering was performed on shared nearest neighbor graph built on the first 30 principal components.

CAR+ and CAR-T cells in CD4/CD8-enriched apheresis, CSF and cyst fluid samples were defined by the presence and absence of GD2-CAR mRNA, respectively. CAR+ T cells in the CAR product samples were defined by either expression of GD2-CAR mRNA or in the CAR+ sorted samples, while CAR-T cells were defined by both absence of GD2-CAR mRNA and in the CAR-sorted samples. Absolute cell numbers in the CSF were calculated using white blood cell counts and CSF lymphocyte percentages obtained from clinical CSF analyses. T cell clonotypes were identified by highly similar (with above 95% sequence similarities) CDR3 nucleotide sequences in both TRA and TRB chains. TRUST4^25^ pipeline was applied to reconstruct BCR sequences from the scRNA-seq dataset. BCR clonotypes were defined by exact match in CDR3 amino acid sequence. Clonal diversity and overlap across timepoints were computed using custom R and python scripts.

### cNMF analysis of T cell programs

The log-normalized count matrix of T cells from CD4/CD8-enriched apheresis, CSF and cyst fluid samples subsetted for the top 2,000 highly variable genes was used as the input for cNMF v.1.6.0^60^. For the cNMF ‘prepare’ step, we performed factorization over K ranges from 10 to 25 and for 500 iterations. K = 14 was chosen based on silhouette score for the cNMF ‘consensus’ step with 10 components and at local-density-threshold value of 0.01. We annotated each program on the final gene_spectra output of cNMF by comparing the top 20 genes with previously published gene sets and known marker genes. The usages matric from cNMF analysis was visualized as FeaturePlot in Seurat. The average program usage score for each sample was used for analyzing associations between T cell cNMF gene programs and clinical parameters.

### Human Anti-CART antibody (HACA) detection by flow cytometry

GD2-CAR-expressing Jurkat cells were used as bait to detect circulating human antibodies against CAR T cells in the plasma and CSF of patients treated with GD2-CAR T cells. Following incubation with patient samples, bound human anti-CAR IgG was detected using Alexa Fluor 488-conjugated anti-human IgG (Thermo Fisher Scientific). Data acquisition was performed on a Cytek Aurora flow cytometer. The percentage of IgG-positive Jurkat cells was used as the primary readout. Signals were normalized to 1A7 anti-GD2-CAR idiotype-stained Jurkat control cells, with the 1A7-positive population defined as 100%. Based on empirical optimization, a 60-70% positive range in the 1A7 control was selected to ensure an appropriate dynamic range for detection. Results are presented as percent positivity relative to the normalized control.

We analyzed the relation between plasma GD2-CAR T cells (Log transformed after adding 1) and HACA using a linear mixed effect model that includes a random intercept (accounting for patient-to-patient variability and within-patient correlation) and two fixed effects: days since first infusion and plasma HACA levels. Only observations after first infusion were considered. AIC and BIC criteria indicated this model to be preferable to one that included only a fixed effect for days, and models that considered a random effect for HACA or a random effect for days. We relied on the lmer function from the lme4 package in R. To compare models, we used maximum likelihood estimation (RMLE=FALSE), and we used restricted maximum likelihood for the final estimation.

### Purification of recombinant GD2-CAR for ELISA

DNA encoding the GD2-CAR (14.G2a) scFv-flag-his was ordered from IDT and cloned into the pAdd2 backbone with EcoRI/XhoI. GD2 CAR was expressed as soluble protein with the Expi293 expression system (Gibco, A14635) and purified with Nickel-NTA Agarose Resin (GoldBio, H-350-5).

### HACA detection by ELISA

Two different ELISAs were performed: 1) Indirect ELISA for the quantification of serum HACA in Pt06 and Pt09 with Pt09 performed in replicate, and 2) Fc-sandwich ELISA to measure the relative expression of recombinant HACA. For all ELISAs, clear, flat-bottom 96w plates (Thermo, 439454) were incubated with 50uL of target protein in PBS overnight, blocked with 3% non-fat milk in PBS for 2hr at room temperature, washed five times between each step with 400 uL PBS + 0.2% Tween-20 (405TS microplate washer, BioTek), bound with reagents diluted in 50 uL PBS + 3% non-fat milk + 0.2% Tween-20, and detected on a microplate reader (Synergy H1, BioTek) after 7 min incubation at with substrate solution (1-Step Ultra TMB, Thermo Fisher, 34028) and addition of 1 M sulfuric acid to stop the solution.

For ELISA #1, purified recombinant 14.G2a-flag-his was coated at 2 ug/mL. Each timepoint of Pt06 and Pt09 serum was diluted 1:1000 and serial diluted 3-fold to the bottom of the plate (A-H), and incubated for 1 hr at room temperature, followed by another 1h incubation with Mouse anti-human IgG Fc-HRP (Southern Biotech, 9042-05) at 1:5000 dilution. To generate the standard curve we utilized anti-14.G2a idiotype^57^, 1A7, which was diluted to 1.6nM and serial diluted 5-fold 8 times, or 100pM and serial diluted 2-fold 8 times. Goat anti-mouse IgG Fc-HRP (Southern Biotech, 1013-05) was diluted 1:10,000, added to every well, and incubated for 1hr at room temperature for detection of 1A7. For ELISA #2 rabbit anti-human IgG (H+L) (Novus Biologicals, NB120-6715) was coated at 2ug/mL. Recombinant HACA supernatant was diluted 1:10, added to each well, and incubated for 1hr at room temperature. Mouse anti-human IgG Fc-HRP (Southern Biotech, 9042-05) was added every well diluted 1:5000 and incubated for 1hr at room temperature.

A single OD450 from each sample that fell within the dynamic range of the concurrent standard curve was taken and used to interpolate a concentration using an unweighted five-parameter logistic (5PL) regression model in GraphPad Prism 10. Each interpolated concentration was subsequently multiplied by the dilution factor to obtain the serum HACA concentration. The analytical limit of detection (LOD) and limit of quantification (LOQ) for the HACA ELISA were established at 0.12 ng/mL and 0.15 ng/mL, respectively.

### IncuCyte Assay

Cocultures of GD2 CAR-T cells and Nalm6-GD2-GFP cells were setup at multiple effector to tumor (E:T) ratios all in RPMI1640 supplemented with 10% FBS, 10mM HEPES, 200mM L-glutamine, 100 U ml^-1^ penicillin and 100 ug ml^-1^ streptomycin. A total of 4 x 10^4^, 1 x 10^4^, and 0.5 x 10^4^ effector cells were cocultured with 4 x 10^4^ tumor cells to create 1:1, 1:4, and 1:8 E:T ratios, respectively. Pt09 high-HACA (Pt09 11_ICV_Late) serum was diluted 1:1000 in RPMI and added to the T cells concurrently with tumor cells. Pt09 serum from a pre-infusion draw was set up the same as previously described only for the 1:1 E:T ratio as a HACA-negative serum control. Cocultures without any patient serum were also set up for each E:T ratio. Tumor fluorescence was monitored every 3 hour with a x10 objective using the IncuCyte S3 Live-Cell Analysis System (Sartorius), housed in a cell culture incubator at 37 °C and 5% CO2, set to take 4 images per well at each timepoint. The total integrated GFP intensity was quantified using the IncuCyte basic analyzer software feature (IncuCyte S3 v.2019B Rev2; Sartorius). Data were normalized to the first timepoint and plotted as the fold change in tumor fluorescence over time. Coculture experiments were set-up using day 9 T cells.

### Recombinant HACA expression

The pcDNA3 vector with signal peptide MEWSWVFLFFLSVTTGVHS was used for antibody expression. DNA encoding the variable heavy and variable light chains for the patient derived HACAs was ordered from IDT. Variable heavy chains were cloned into an AgeI/ApaI-digested pcDNA3 vector containing hIgG1 CH1-hinge-CH2-CH3 using Gibson assembly. Variable light chains were cloned into either an AgeI/Bsu36I-digested (hIGL lambda) or AgeI/BsiWI-digested (hIGL kappa) pcDNA3 vector containing the corresponding hIGL using Gibson assembly. Plasmids were co-transfected into Expi293 cells (Thermo Fisher Scientific) at a 1:1 ratio of heavy chain:light chain using ExpiFectamine according to manufacturer’s instructions. Then, after 5 days of transfection, the supernatant was collected by centrifugation, sterile-filtered, and stored at 4°C.

### Flow cytometry analysis of recombinant HACA binding to T cells

Cells were washed with FACS buffer (2% FBS in PBS) before staining (100,000 cells/condition). Cells were stained with 100uL of recombinant antibody supernatant, patient serum, or control antibody for 45 minutes at 4°C before washing once with FACS buffer and staining with anti-human IgG-AF647 (Southern Biotech, 2040-31) for 20 minutes at 4°C. After staining, cells were washed once with FACS buffer and analyzed on the Thermo AttuneTM NxT system. AttuneTM NxT software (v7.1, Thermo) was used for data collection and FlowJo software (v10.10.0; BD) was used for data analysis.

### Peptide preparation for T cell assays

The full ORF (FKBP-iCasp9-T2A-ERsignal-VL_14g2a_-VH_14g2a_-whitlow-CD8-41BB-CD3z) of the GD2-encoding retroviral vector used for manufacturing the CAR T cell product was translated into a peptide library of 150 peptides that were synthesized by GenScript at a purity of ≥75%. We chose a design of 18 amino acid long peptides overlapping by 12 residues (off-set 6) to achieve a compromise between typical lengths of CD8^+^ and CD4^+^ T cell epitopes. Lyophilized peptides were reconstituted at 100 mg mL^-1^ in DMSO (Sigma, Cat#D8418). Individual peptides were pooled 1:1000 in RPMI 1640 (Life Technologies, Cat#31870-025), creating a master pool with each peptide at 100 µg mL^-1^ in RPMI 1640 15% DMSO. We further created 16 overlapping peptide pools with each peptide at 500 µg mL^-1^ in RPMI 1640 15% DMSO according to a 3-D-matrix design^61^ (Supplementary Table 6). Each pool contained up to 30 peptides and each peptide occurred in three pools, while the pool ID combination for each peptide was unique (5 x 5 x 6 = 150). Pools were aliquoted and stored at-80°C.

### Antigen-specific expansion of T cells

To compensate for the low frequency of antigen-specific T cells even among non-naïve populations, we adjusted established protocols^61^ (and Koch *et al.* in preparation) for antigen-specific in-vitro expansion by peptide stimulation to our needs. Frozen PBMCs were thawed in a 37°C warm water bath and washed with warm RPMI 1640 supplemented with 5 mM HEPES (Roth, Cat#HN77.4), 1% Penicillin-Streptomycin-Glutamine (Life Technologies, Cat#10378016), and 10% FCS from Life Technologies (RPMI/FCS). Cells were rested overnight in RPMI/FCS in a humidified incubator at 37°C, 5% CO_2_. Cells were then harvested, washed with RPMI 1640 supplemented with 5 mM HEPES, 1% Penicillin-Streptomycin-Glutamine, plus 5% human serum (BIOIVT, Cat#HUMANABSRMP-HI-1) (RPMI/HS), and count manually in a Neubauer Chamber with Trypan Blue counterstaining. App. 1 ×10^6^ PBMCs were pulsed with the master peptide pool at 5 µg mL^-1^ per peptide in RPMI/HS on a vertical tube roller for 1h at RT. Excess of peptide was washed of and pulsed cells were returned to app. 5 ×10^6^ PBMCs from the same sample at a final concentration of 2 ×10^6^ cells mL^-1^ in a 12-well-plate. Cells were incubated in RPMI/HS supplemented with 50 U mL^-1^ IL-2 (Peprotech, Cat#200-02) and 25 µg mL^-1^ IL-7 (Peprotech, Cat#200-07) at 37°C, 5% CO_2_ and RPMI/HS plus IL-2 was refreshed every 3 days.

### T cell re-stimulation with peptide library

After 8 full days of expansion, cells were harvested, washed, and rested for 4h in RPMI/HS. Cells were split into 20 subsamples on a U-bottom 96-well-plate, each containing RPMI/HS supplemented with Brefeldin A (1:1000; BD Biosciences, Cat#555029), Monensin (1:1500; BD Biosciences, Cat#554724), and CD107a FITC (1:200; all antibodies are listed in Supplementary Table S7) at a final volume of 200 µL. Each of the 16 3-D-matrix pools were added at 1 µg mL^-1^ per peptide to one of the subsamples. The positive control samples received CEFX-peptide pool (JPT Peptides, PM-CEFX-1) at app. 1 µg mL^-1^ per peptide and PMA+Ionomycin (1:500; Cell Stimulation Cocktail, eBioscience, Cat#P1585), respectively. Two samples supplemented with peptide solvent only (RPMI 1640 15% DMSO) at equal concentration to peptide stimulation served as negative control replicates. Cells were incubated over night at 37°C, 5% CO_2_. Culture was terminated and cells were transferred to a 96-well V-bottom plate on ice/at 4°C. After centrifugation at 500 g for 3 min, RPMI/HS was discarded, and cells were washed in PBS. For live/dead-discrimination, cells were incubated with Live/Dead Blue (Invitrogen, Cat#L23105) in PBS (1:1000) for 20 min on ice under light protection. Cells were then washed with FACS buffer (PBS 0.5% BSA) and stained with fluorophore-conjugated antibodies CD45RA BUV496 (1:100), CCR7 PerCP (1:100), CD14 PE-Dazzle (1:100), CD19 PE-Dazzle (1:100), and CD56 PE-Dazzle (1:100) in 50 µL FACS buffer for 30 min on ice. After washing with FACS buffer, cells were fixed and then washed with Cytofix/Cytoperm buffer solutions (BD Biosciences, Cat#554714) followed by staining with CD3 BUV805 (1:100), CD8α BUV615 (1:400), CD4 BUV395 (1:100), CD40L BV421 (1:50), 4-1BB BV480 (1:100), anti-IFN-γ PE (1:40), anti-IL-2 PE-Cy7 (1:40), anti-IL-4 BV605 (1:40), anti-IL-17a BV785 (1:40), anti-IL-21 APC (1:40), anti-TNF APC-Fire810 (1:40), and anti-IL-10 BB700 (1:40) in 50 µL Perm/Wash supplemented with Brilliant Stain Buffer Plus (1:10; BD Biosciences, Cat#566385). CD3, CD4, and CD8α were stained intracellularly to compensate for activation-induced internalization. The surface activation markers 41BB and CD40L were also stained intracellularly to compensate for the impaired protein trafficking induced by Golgi-transport inhibitors^62^. After washing with PermWash and once with FACS buffer, cells were resuspended in 200 µL FACS buffer and analyzed under identical parameters on a Cytek Aurora (Cytek Biosciences) equipped with 5 lasers (355, 405, 488, 561, 640 nm) and 64 detectors. Unmixed data was analyzed in FlowJo. Unmixing was performed based on single color-stained Beads (BD Biosciences, Cat#569991) or PBMCs (Live/Dead Blue) that were processed equally to the samples (reference controls). Unmixed data was analyzed in FlowJo (BD, FlowJo 10.10.1). The percentage of cells within the CD4^+^ or CD8^+^ T cell population of a peptide-stimulated sample was normalized by subtracting mean cytokine^+^ percentage of its corresponding DMSO control samples.

### Statistical analysis for T cell response

Flow cytometry data and associated metadata were organized as a tidy data frame (Supplementary Table 8) and analyzed in Python. Low-quality samples, including those with <1,000 acquired T cells, were excluded. For each peptide-stimulated sample, the percentage of cytokine-positive CD4⁺ or CD8⁺ T cells was normalized by subtracting the mean of the corresponding DMSO control. Response magnitudes, representing the overall response to the CAR transgene, were calculated by summing up normalized responses across peptide pools and dividing by three. Group-wise comparisons (with or without stratification by time relative to HACA emergence) were performed using linear mixed-effects models. Normalized responses were modeled as a function of treatment (CAR-treated versus healthy controls), with patient included as a random effect to account for repeated measurements and peptide pool (restimulation) included as an additional variance component. Models were fitted using restricted maximum likelihood. Fixed-effect estimates (β) and corresponding 95% confidence intervals were visualized as forest plots, with statistical significance derived from the model. For pool-level analyses (including per-patient analyses and epitope mapping), samples were required to contain ≥3,000 T cells and ≥5 cytokine-positive events. Antigen-specific responses were defined as those exceeding both a two-fold increase over the sample-specific DMSO control and a control-derived threshold (mean + 3× standard deviation of healthy controls). To assess whether individual patient responses exceeded those observed in healthy controls, one-sided permutation tests were performed. For each patient, the observed statistic was defined as the mean response across peptide pools. A null distribution was generated by resampling (10,000 iterations, with replacement) from pooled healthy control responses, matched to the number of pools per patient. Empirical p-values were calculated as the proportion of permuted means greater than or equal to the observed mean and adjusted for multiple testing across patients using Bonferroni correction. For epitope mapping, true responses were evaluated across three orthogonal peptide pool dimensions (pools 1–5, 6–10, and 11–16). Assuming equal distribution of antigen-specific T cells across subsamples prior to restimulation, concordant responses of similar magnitude and phenotype across all three dimensions were used to infer the corresponding immunogenic peptide. Only unambiguous triplet intersections were considered. Plots were generated using Matplotlib and Seaborn libraries in Python.

## Statistical analysis

All computations were performed in R or python. No data were excluded except for established single-cell QC filters.

## Data availability

Single-cell RNA sequencing data will be available upon publication.

## Code availability

The scripts and codes used to generate all the data in the study will be available on GitHub.

## Data Availability

All data produced in the present study are available upon reasonable request to the authors

## Acknowledgements

This work was supported by NIH Grants: R35CA283888 (C.L.M.), OT2OD038101 (C.L.M., Z.G.), 2P30CA124435-16, the California Institute for Regenerative Medicine (CLIN2-12595, C.L.M., M.M.), the Virginia and D.K. Ludwig Fund for Cancer Research (C.L.M.). C.L.M. and S.A.Y-H are members of the Parker Institute for Cancer Immunotherapy, which supports the Stanford University Cancer Immunotherapy Program. C.L.M., Y.C., S.R. and E.S. are Weill West Coast Cancer Hub Investigators. M.R.A.K. was supported by a Walter Benjamin Fellowship from the DFG (project number 551714776). Tyler E. Miller and Edward Estrada from Case Western assisted with cNMF and provided feedback on our analyses. Research reported in this publication was supported by the National Center For Advancing Translational Sciences of the National Institutes of Health under Award Number UM1TR004921. The content is solely the responsibility of the authors and does not necessarily represent the official views of the National Institutes of Health.

## Author contributions

M.M. is principal investigator of the trial and C.L.M. is the IND holder. M.M., R.M., J.M., K.W.S., S.R. planned, designed and/or wrote the clinical trial and protocol amendments. K.W.S., J.M., R.M., J.G., L.P., C.C, S.P., K.D., J.M., C.E., A.J., A.D., S.R. participated in clinical care. E.E., C.B., M.F., M.K., monitoring patient toxicity, participated in collection and processing of patient samples and provided regulatory oversight. S.P. created data infrastructure that was utilized. K.R., M.R.A.K., C.P., M.D., B.S., A.K., S.R.S., M.D., Y.W.H., Z.E., N.I., C.S. generated data. Y.C., M.R.A.K., K.C.Y.T., Z.G., K.M. S.Y.H., E.S., C.L.M., S.R., analyzed data. B.S., C.L.M., S.R., S.H., J.C., oversaw data collection. R.T., S.F. oversaw product manufacturing. Y.C., K.R., M.R.A.K., S.R., C.L.M., generated figures and wrote the manuscript. M.M., S.R., and C.L.M. supervised all aspects of the work.

## Competing interests

C.L.M., M.M., R.J.M. are coinventors on a patent related to this work (U.S. Patent <u>17/815,056</u> issued to Stanford University). C.L.M. holds equity in Link Cell Therapies and Ensoma, which are developing CAR-based therapies and consults for Link, Immatics, Ensoma, Astra-Zeneca, Moderna, Kite Pharma, Nektar and Red Tree Capital. E.S. holds equity in Lyell Immunopharma and consults for Lepton Pharmaceuticals.

## Extended Figures

**Extended Data Fig. 1: Cell type annotation and composition in the single-cell atlas**

**a**, Outline for clinical trial design. Retreatment was offered to patients who experienced radiographic or neurologic benefit as defined in our previous publications^15,16^. Repeated ICV infusions were allowed no sooner than 4 weeks after prior treatment, but infusion timelines were variable between patients in Arm A of the study. Abbreviations: D-4 to D-2, Day-4 to-2, D0: Day 0; D28, Day 28; LD, lymphodepleting chemotherapy; IV, intravenous; ICV, intracerebroventricular.

**b**, Collection of 181 samples from 13 DMG patients during GD2-CAR T cell therapy for single-cell sequencing. The color represents the total number of cells collected at the indicated timepoints. Note that there could be multiple apheresis and products from the same patient, or multiple CSF and cyst fluid samples collected within the same timepoint but at different days or time of the day, and in such cases the cell numbers shown in the block represent the combined cell numbers. IV-early and IV-late refer to day 1-14 and day 15+ after IV infusion. ICV-early, ICV-mid and ICV-late refer to day 1-5, day 6-14 and day 15+ after ICV infusions, respectively.

**c**, UMAP visualization of clustering, cell cycle phase prediction and automatic cell type annotation results.

**d**, UMAP visualization of representative myeloid cell type marker expression and H3K27M+cells with at least 3 UMIs carrying the mutant allele.

**Extended Data Fig. 2: Lymphocyte compositions across samples**

**a**, Pie charts depicting the fractions of CAR+ T cells (maroon) and CAR^-^ T cells (grey) across sample types and timepoints (left), and within each T cell subtype in post-treatment CSF (right).

**b-c**, Fraction of lymphocyte cell types across CD4/CD8-enriched apheresis and CAR products (b), and in CSF and cyst fluid samples (c). Tumor and myeloid cell types were summed and annotated as other cell types.

**d**, Proportions of CAR^+^ and CAR^-^ T cell subtypes across sample types and timepoints. Each sample is represented as a scatter and colored according to patient ID. The black bar marks the median. The vertical lines mark different infusion cycles. IV-early and IV-late referred to day 1-14 and day 15+ after IV infusion. ICV-early, ICV-mid and ICV-late referred to day 1-5, day 6-14 and day 15+ after ICV infusions, respectively.

**e**, The log2(CD4:CD8) ratio in CAR+ T cells (top), CAR-T cells (middle), and the proportions of Tregs amongst all CD4+ T cells (bottom) across sample types and timepoints.

**f**, Proportions of B cells, plasmablast and NK cells across sample types and timepoints.

**Extended Data Fig. 3: Lymphocyte dynamics across infusion cycles**

**a**, Estimated absolute number of lymphocyte cell subtypes in the CSF in each patient, based on cell fractions in scRNA-seq adjusted by clinical CSF white blood cell count and lymphocyte fraction. Note that 5 patients (Pt02, Pt04, Pt11, Pt12, Pt13) did not have clinical CSF white blood cell counts available and thus were not included in this analysis.

**b**, Dynamics of cell fractions in the CSF at baseline and at each infusion cycle of the GD2-CAR T cell therapy. Fractions were calculated by dividing the number of cells from each cell type by the total number of cells in the sample.

**c-d**, Volcano plots comparing the fractions (c) and absolute cell number (d) in samples from infusions 1-5 versus infusions 6-17. P-values were calculated using the Wilcoxon rank-sum test and corrected for multiple comparisons.

**Extended Data Fig. 4: TCR repertoire across patients, samples and treatment timelines**

**a-c**, Number of TCR clonotypes in all T cells (a), CAR+ T cells (b) and CAR-T cells (c) in each patient. Color represents number of clonotypes detected uniquely in one sample type or across sample types.

**d**, Number of TCR clonotypes shared between CAR+ (top) and CAR-(bottom) T cells in CAR product, CSF, and all T cells from CD4/CD8-enriched apheresis. The CAR+ versus CAR-status in CD4/CD8-enriched apheresis were omitted as the fraction of CAR+ cells in apheresis was low.

**e**, Number of TCR clonotypes shared between CD4/CD8-enriched apheresis and CAR products. The patients were grouped by whether one or more products were generated and whether these products originated from one or multiple apheresis collections. The colored bars indicated the patient identity of each apheresis and product sample. The matched apheresis and CAR products were highlighted with red rectangles.

**f**, Clonality of TCR repertoire in CD4/CD8-enriched apheresis samples. Each sample is represented as a scatter and colored according to patient ID. The boxplot shows the distributions across samples, with the center line presenting the median and box representing the 1^st^ and 3^rd^ quantiles.

**g**, Clonality of CAR+ (circle and solid line) and CAR-(triangle and dashed line) TCR repertoire in CAR product.

**Extended Data Fig. 5: TCR repertoire dynamics in CAR+ T cells**

**a**, TCR clonotype frequency in CAR+ T cells across sample types. Only CAR+ T cells with defined TCR clonotypes were colored, while all other cells in the sample were grey and in the background.

**b**, Pairwise TCR repertoire similarity between samples from the same patient by the Morisita-Horn index.

**c**, Clonotype fractions of top10 CAR+ T cell clonotypes in Pt06, Pt07 and Pt09. Each color represented one TCR clonotype. The triangles on top marked different products in the patient, and the bar marked the product used in different infusion cycles.

**Extended Data Fig. 6: TCR repertoire dynamics in CAR-T cells**

**a**, TCR clonotype frequency in CAR+ T cells across sample types. Only CAR-T cells with defined TCR clonotypes were colored, while all other cells in the sample were grey and in the background.

**b**, Clonality index of CAR-CD8+ T cells (top), CAR-CD4+ T cells (middle) and CAR-Tregs (bottom) in CSF samples with at least 50 cells. Each sample is represented as a scatter and colored according to patient ID. The vertical lines mark different infusion cycles. IV-early and IV-late referred to day 1-14 and day 15+ after IV infusion. ICV-early, ICV-mid and ICV-late referred to day 1-5, day 6-14 and day 15+ after ICV infusions, respectively.

**c**, Fractions of CAR-Tregs with selected highly expanded clonotypes out of the CAR-Treg TCR repertoire.

**d**, Volcano plot highlighting top differential expressed genes (DEG) in CAR-Tregs that were highly expanded (with ≥5 CAR-Tregs) versus less expanded. X-axis represented the log2(fold-change) of average gene expression level in hyper-expanded versus less expanded CAR-Treg clonotypes, and Y-axis represented the-log10(adjusted P-values) by Wilcoxon rank-sum test.

**Extended Data Fig. 7: Characterization of cNMF programs of T cells in CSF and apheresis**

**a**, UMAP embedding of T cells from CSF and CD4/CD8-enriched apheresis, colored by T cell subtypes, sample type, cell cycle phase, CAR mRNA expression, CAR-and CAR+ clonotype frequency, respectively.

**b**, Hierarchical clustering of CSF and CD4/CD8-enriched apheresis samples based on mean scores of cNMF program activities. Top tracks annotated the sample information and clonality scores of CAR-and CAR+ T cells.

**c**, UMAP visualization of selected cNMF programs.

**d**, Average score for program 9 activity in T cells from ICV-early CSF samples from patients with sustained (>12m) versus intermediate (5-12m) response to GD2-CAR T cell therapy. P-value was calculated by Wilcoxon rank-sum test.

**Extended Data Fig. 8: Detection of adaptive cellular immunity against GD2-CAR in patient PBMC**

**a**, Experimental setup for ex-vivo stimulation-restimulation of PBMCs with CAR-derived peptides and subsequent analysis for T cell responses. 5 x10^6^ cryo-conserved PBMCs were thawed and rested overnight. 20% of the cells were then pulsed with the whole CAR peptide library for 2h and mixed with the remaining sample to facilitate expansion of antigen-specific T cells. After 8 days of cell culture in the presence of IL-2 and IL-7, cells were washed and rested for 6h. 20 subsamples were subsequently created and each restimulated overnight with one of 16 three-dimensional-matrix peptide pools or positive and negative controls. Activation-induced marker expression and cytokine secretion was measured via flow cytometry.

**b**, Gating strategy to identify CD4⁺ and CD8⁺ T cells in PBMC samples.

**c**, Peptide library design and pooling strategy according to a three-dimensional matrix design. The full translated amino acid sequence as encoded by the GD2 CAR-vector was translated into 150 peptides (one omitted) with a length of 18 amino acids, that overlapped by 12 amino acids (off-set 6). Synthesized peptides were dissolved in DMSO and added to one of the peptide pools 1-5 (X dimension), one of the peptide pools 6-10 (Y dimension), and one of the peptide pools 11-16 (Z dimension). Each peptide was therefore defined by a unique set of pools in which it occurred.

**d**, Representative flow cytometry plots displaying IIL-4, IL-17A, and IL-10 expression alongside the IFN-ɣ and TNFα signal in CD4^+^ T cells.

**e**, Representative flow cytometry plots displaying IFN-γ and TNFα secretion of CD8^+^ T cells.

**Extended Data Fig. 9: Permutation testing for defining anti-CAR cellular immunity.**

Each plot compares the observed mean response across 16 peptide pools for the indicated patient and parameter to a null distribution generated from permuted responses of n = 4 healthy donors. Statistical significance is indicated by a red line. A response was considered positive if statistically significant plus if it exceeded a twofold increase over the patient-specific DMSO control and met minimum event thresholds (≥3,000 total T cells and ≥5 cytokine-positive events).

**Extended Data Fig. 10: HACA detection in plasma and CSF across patients**

**a**, Longitudinal profiling of GD2-CAR copies (left axis) and human anti-CAR antibody (HACA) ratio (right axis) in plasma and CSF by flow cytometry assay. Response per infusion was marked on top of the figures.

**b**, Area under the curve (AUC) analysis representing tumor cell survival over the first 24 hours of coculture between GD2-CAR T cells and Nalm6-GD2 target cells showed in **Fig. 5f**. Data are presented as mean ± SEM of n=3 replicates. Statistical significance was determined by one-way ANOVA followed by Tukey’s multiple comparisons test. ns, not significant; **P < 0.01; ***P < 0.001; ****P < 0.0001.

**Extended Data Fig. 11: Re-expression of antibodies reconstructed from scRNA-seq**

**a**, Fraction of immunoglobin isotypes in the TRUST4-reconstructed, expanded BCR clonotypes (with at least 3 B cells per clonotype) in Pt06, Pt07 and Pt09.

**b**, Expression of recombinant antibodies in the supernatant measured by A450 absorbance. The serum samples of Pt09 at pre-and on-treatment timepoints were included for comparisons of antibody expression levels.

**c**, Longitudinal immune and clinical profiles of patients Pt03 and Pt04. The first four tracks captured anti-CAR T cell immune responses measured in peripheral blood. Specifically, the percentages of CD40L+41BB+ and IFNγ+TNF+ cells among CD4+ T cells (tracks 1-2, orange bars) and CD8+ T cells (tracks 3-4, blue bars) were quantified at each timepoint by intracellular cytokine stimulation assay. Plasma HACAs (track 5, red) and CSF HACAs (track 7, red) levels were measured by flow cytometry. Plasma CAR T cell copy number per 100ng (track 6, black) were measured by qPCR. Percentages of B cells (track 8, yellow) and CAR+ T cells among total cells in CSF (track 9, black) were derived from scRNA-seq data. The clinical timeline at the bottom of each column indicates the treatment course, with colored segments denoting response per infusion cycle. The dashed vertical red line marks the approximate onset of HACA in plasma.

## Notes

### Competing Interest Statement

C.L.M., M.M., R.J.M. are coinventors on a patent related to this work (U.S. Patent 17/815,056 issued to Stanford University). C.L.M. holds equity in Link Cell Therapies and Ensoma, which are developing CAR-based therapies and consults for Link, Immatics, Ensoma, Astra-Zeneca, Moderna, Kite Pharma, Nektar and Red Tree Capital. E.S. holds equity in Lyell Immunopharma and consults for Lepton Pharmaceuticals.

### Author Declarations

IRB of Stanford University gave ethical approval for this work

## References

1. Majzner, R. G. et al. GD2-CAR T cell therapy for H3K27M-mutated diffuse midline gliomas. Nature 603, 934–941 (2022).

2. Monje, M. et al. Intravenous and intracranial GD2-CAR T cells for H3K27M+ diffuse midline gliomas. Nature 637, 708–715 (2025).

3. Hansen, D. K. et al. Comparison of Standard-of-Care Idecabtagene Vicleucel and Ciltacabtagene Autoleucel in Relapsed/Refractory Multiple Myeloma. J Clin Oncol 43, 1597–1609 (2025).

4. Berdeja, J. G. et al. Ciltacabtagene autoleucel, a B-cell maturation antigen-directed chimeric antigen receptor T-cell therapy in patients with relapsed or refractory multiple myeloma (CARTITUDE-1): a phase 1b/2 open-label study. Lancet 398, 314–324 (2021).

5. Frank, M. J. et al. CD22-directed CAR T-cell therapy for large B-cell lymphomas progressing after CD19-directed CAR T-cell therapy: a dose-finding phase 1 study. Lancet 404, 353–363 (2024).

6. Neelapu, S. S. et al. Axicabtagene Ciloleucel CAR T-Cell Therapy in Refractory Large B-Cell Lymphoma. N Engl J Med 377, 2531–2544 (2017).

7. Jain, M. D. et al. Five-Year Follow-Up of Standard-of-Care Axicabtagene Ciloleucel for Large B-Cell Lymphoma: Results From the US Lymphoma CAR T Consortium. J Clin Oncol 42, 3581–3592 (2024).

8. Schultz, L. M. et al. Disease Burden Affects Outcomes in Pediatric and Young Adult B-Cell Lymphoblastic Leukemia After Commercial Tisagenlecleucel: A Pediatric Real-World Chimeric Antigen Receptor Consortium Report. J Clin Oncol 40, 945–955 (2022).

9. Maude, S. L. et al. Tisagenlecleucel in Children and Young Adults with B-Cell Lymphoblastic Leukemia. N Engl J Med 378, 439–448 (2018).

10. Abramson, J. S. et al. Lisocabtagene maraleucel for patients with relapsed or refractory large B-cell lymphomas (TRANSCEND NHL 001): a multicentre seamless design study. Lancet 396, 839–852 (2020).

11. Del Bufalo, F. et al. GD2-CART01 for Relapsed or Refractory High-Risk Neuroblastoma. N Engl J Med 388, 1284–1295 (2023).

12. Qi, C. et al. Claudin-18 isoform 2-specific CAR T-cell therapy (satri-cel) versus treatment of physician’s choice for previously treated advanced gastric or gastro-oesophageal junction cancer (CT041-ST-01): a randomised, open-label, phase 2 trial. Lancet 405, 2049–2060 (2025).

13. Steffin, D. et al. Interleukin-15-armoured GPC3 CAR T cells for patients with solid cancers. Nature 637, 940–946 (2025).

14. Bagley, S. J. et al. Intracerebroventricular bivalent CAR T cells targeting EGFR and IL-13Rα2 in recurrent glioblastoma: a phase 1 trial. Nat Med 1–10 (2025) doi:10.1038/s41591-025-03745-0.

15. Brown, C. E. et al. Locoregional delivery of IL-13Rα2-targeting CAR-T cells in recurrent high-grade glioma: a phase 1 trial. Nat Med 30, 1001–1012 (2024).

16. Choi, B. D. et al. Intraventricular CARv3-TEAM-E T Cells in Recurrent Glioblastoma. New England Journal of Medicine 390, 1290–1298 (2024).

17. Aran, D. et al. Reference-based analysis of lung single-cell sequencing reveals a transitional profibrotic macrophage. Nature Immunology 20, 163–172 (2019).

18. Hao, Y. et al. Integrated analysis of multimodal single-cell data. Cell 184, 3573–3587.e29 (2021).

19. Walker, A. J. et al. Tumor Antigen and Receptor Densities Regulate Efficacy of a Chimeric Antigen Receptor Targeting Anaplastic Lymphoma Kinase. Mol Ther 25, 2189–2201 (2017).

20. Li, W. et al. Chimeric Antigen Receptor Designed to Prevent Ubiquitination and Downregulation Showed Durable Antitumor Efficacy. Immunity 53, 456–470.e6 (2020).

21. van Gisbergen, K. P. J. M., Zens, K. D. & Münz, C. T-cell memory in tissues. European Journal of Immunology 51, 1310–1324 (2021).

22. Parry, E. M. et al. ZNF683 marks a CD8+ T cell population associated with anti-tumor immunity following anti-PD-1 therapy for Richter syndrome. Cancer Cell 41, 1803–1816.e8 (2023).

23. Good, C. R. et al. An NK-like CAR T cell transition in CAR T cell dysfunction. Cell 184, 6081–6100.e26 (2021).

24. Pattekar, A. et al. Norovirus-Specific CD8+ T Cell Responses in Human Blood and Tissues. Cellular and Molecular Gastroenterology and Hepatology 11, 1267–1289 (2021).

25. Song, L. et al. TRUST4: immune repertoire reconstruction from bulk and single-cell RNA-seq data. Nat Methods 18, 627–630 (2021).

26. Cyster, J. G. & Allen, C. D. C. B Cell Responses: Cell Interaction Dynamics and Decisions. Cell 177, 524–540 (2019).

27. Howard, E. L., Goens, M. M., Susta, L., Patel, A. & Wootton, S. K. Anti-Drug Antibody Response to Therapeutic Antibodies and Potential Mitigation Strategies. Biomedicines 13, 299 (2025).

28. Lee, D. W. et al. T cells expressing CD19 chimeric antigen receptors for acute lymphoblastic leukaemia in children and young adults: a phase 1 dose-escalation trial. Lancet 385, 517–528 (2015).

29. Turtle, C. J. et al. CD19 CAR-T cells of defined CD4+:CD8+ composition in adult B cell ALL patients. J Clin Invest 126, 2123–2138 (2016).

30. Wagner, D. L. et al. Immunogenicity of CAR T cells in cancer therapy. Nat Rev Clin Oncol 18, 379–393 (2021).

31. Cappell, K. M. & Kochenderfer, J. N. Long-term outcomes following CAR T cell therapy: what we know so far. Nat Rev Clin Oncol 20, 359–371 (2023).

32. Narayan, V. et al. PSMA-targeting TGFβ-insensitive armored CAR T cells in metastatic castration-resistant prostate cancer: a phase 1 trial. Nat Med 28, 724–734 (2022).

33. O’Rourke, D. M. et al. A single dose of peripherally infused EGFRvIII-directed CAR T cells mediates antigen loss and induces adaptive resistance in patients with recurrent glioblastoma. Sci Transl Med 9, eaaa0984 (2017).

34. Adusumilli, P. S. et al. A Phase I Trial of Regional Mesothelin-Targeted CAR T-cell Therapy in Patients with Malignant Pleural Disease, in Combination with the Anti-PD-1 Agent Pembrolizumab. Cancer Discov 11, 2748–2763 (2021).

35. Amini, L. et al. Preparing for CAR T cell therapy: patient selection, bridging therapies and lymphodepletion. Nat Rev Clin Oncol 19, 342–355 (2022).

36. Bot, A. et al. In vivo chimeric antigen receptor (CAR)-T cell therapy. Nat Rev Drug Discov 25, 116–137 (2026).

37. Aalipour, A., Barreiro, A., Garmilla, A. & Birnbaum, M. E. In vivo CAR T cell engineering: design principles and open questions. Trends Cancer S2405–8033(26)00007–5 (2026) doi:10.1016/j.trecan.2026.01.007.

38. Depil, S., Duchateau, P., Grupp, S. A., Mufti, G. & Poirot, L. ‘Off-the-shelf’ allogeneic CAR T cells: development and challenges. Nat Rev Drug Discov 19, 185–199 (2020).

39. Liu, F. et al. Selective HLA knockdown and PD-L1 expression prevent allogeneic CAR-NK cell rejection and enhance safety and anti-tumor responses in xenograft mice. Nat Commun 16, 8809 (2025).

40. Jo, S. et al. Endowing universal CAR T-cell with immune-evasive properties using TALEN-gene editing. Nat Commun 13, 3453 (2022).

41. Degagné, É. et al. High-Specificity CRISPR-Mediated Genome Engineering in Anti-BCMA Allogeneic CAR T Cells Suppresses Allograft Rejection in Preclinical Models. Cancer Immunol Res 12, 462–477 (2024).

42. Gornalusse, G. G. et al. HLA-E-expressing pluripotent stem cells escape allogeneic responses and lysis by NK cells. Nat Biotechnol 35, 765–772 (2017).

43. Deuse, T. et al. Hypoimmunogenic derivatives of induced pluripotent stem cells evade immune rejection in fully immunocompetent allogeneic recipients. Nat Biotechnol 37, 252–258 (2019).

44. Wang, B. et al. Generation of hypoimmunogenic T cells from genetically engineered allogeneic human induced pluripotent stem cells. Nat Biomed Eng 5, 429–440 (2021).

45. Mo, F. et al. Engineered off-the-shelf therapeutic T cells resist host immune rejection. Nat Biotechnol 39, 56–63 (2021).

46. Medawar, P. B. Immunity to homologous grafted skin; the fate of skin homografts transplanted to the brain, to subcutaneous tissue, and to the anterior chamber of the eye. Br J Exp Pathol 29, 58–69 (1948).

47. Smyth, L. C. D. & Kipnis, J. Redefining CNS immune privilege. Nat Rev Immunol 25, 766–775 (2025).

48. Ahn, J. H. et al. Meningeal lymphatic vessels at the skull base drain cerebrospinal fluid. Nature 572, 62–66 (2019).

49. Smyth, L. C. D. et al. Identification of direct connections between the dura and the brain. Nature 627, 165–173 (2024).

50. Fitzpatrick, Z. et al. Venous-plexus-associated lymphoid hubs support meningeal humoral immunity. Nature 628, 612–619 (2024).

51. Theruvath, J. et al. Locoregionally administered B7-H3-targeted CAR T cells for treatment of atypical teratoid/rhabdoid tumors. Nat Med 26, 712–719 (2020).

52. Fitzpatrick, Z. et al. Gut-educated IgA plasma cells defend the meningeal venous sinuses. Nature 587, 472–476 (2020).

53. Magoola, M. & Niazi, S. K. Engineering Universal Cancer Immunity: Non-Tumor-Specific mRNA Vaccines Trigger Epitope Spreading in Cold Tumors. Vaccines (Basel) 13, 970 (2025).

54. Liu, J. et al. Endogenous immune recruitment in glioblastoma CAR T therapy: cytokine, myeloid, and chemokine circuitry. J Neurooncol 177, 50 (2026).

55. Zuo, Y. et al. IL-36γ armored CAR T cells reprogram neutrophils to induce endogenous antitumor immunity. Cancer Cell 0, (2025).

56. Labanieh, L. & Mackall, C. L. CAR immune cells: design principles, resistance and the next generation. Nature 614, 635–648 (2023).

57. Sen, G., Chakraborty, M., Foon, K. A., Reisfeld, R. A. & Bhattacharya-Chatterjee, M. B. Induction of IgG antibodies by an anti-idiotype antibody mimicking disialoganglioside GD2. J Immunother 21, 75–83 (1998).

58. Monaco, G. et al. RNA-Seq Signatures Normalized by mRNA Abundance Allow Absolute Deconvolution of Human Immune Cell Types. Cell Reports 26, 1627–1640.e7 (2019).

59. Huang, X. & Huang, Y. Cellsnp-lite: an efficient tool for genotyping single cells. Bioinformatics 37, 4569–4571 (2021).

60. Kotliar, D. et al. Identifying gene expression programs of cell-type identity and cellular activity with single-cell RNA-Seq. eLife 8, e43803 (2019).

61. Tomov, V. T. et al. Persistent enteric murine norovirus infection is associated with functionally suboptimal virus-specific CD8 T cell responses. J Virol 87, 7015–7031 (2013).

62. Litjens, N. H. R., de Wit, E. A., Baan, C. C. & Betjes, M. G. H. Activation-induced CD137 is a fast assay for identification and multi-parameter flow cytometric analysis of alloreactive T cells. Clin Exp Immunol 174, 179–191 (2013).

