## Supplementary figures and images for "Anti-CAR Immunity Drives Acquired Therapeutic Resistance to GD2-CAR T Cell Therapy in Diffuse Midline Glioma"

### Extended Data Fig. 1

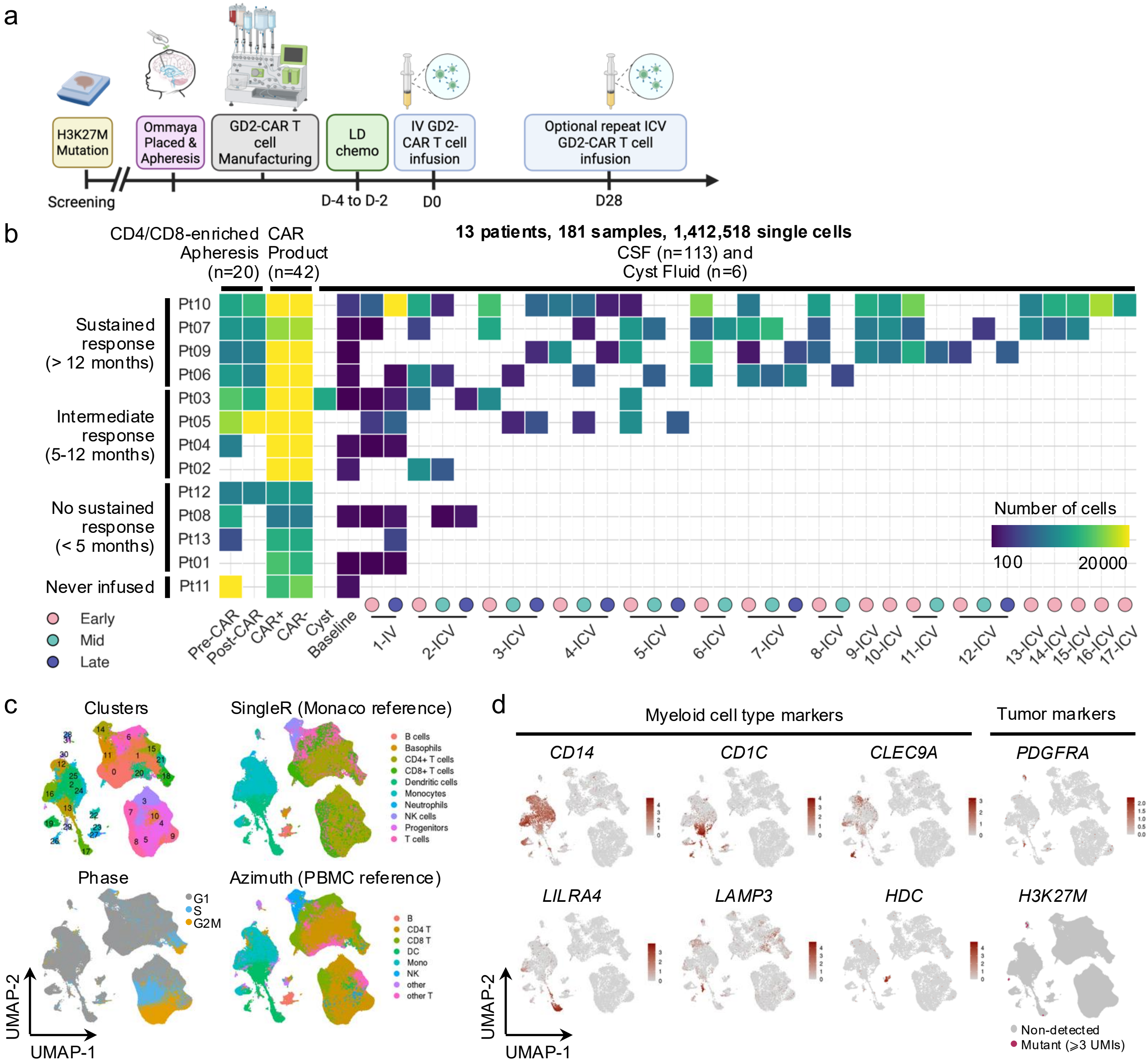

### Extended Data Fig. 2

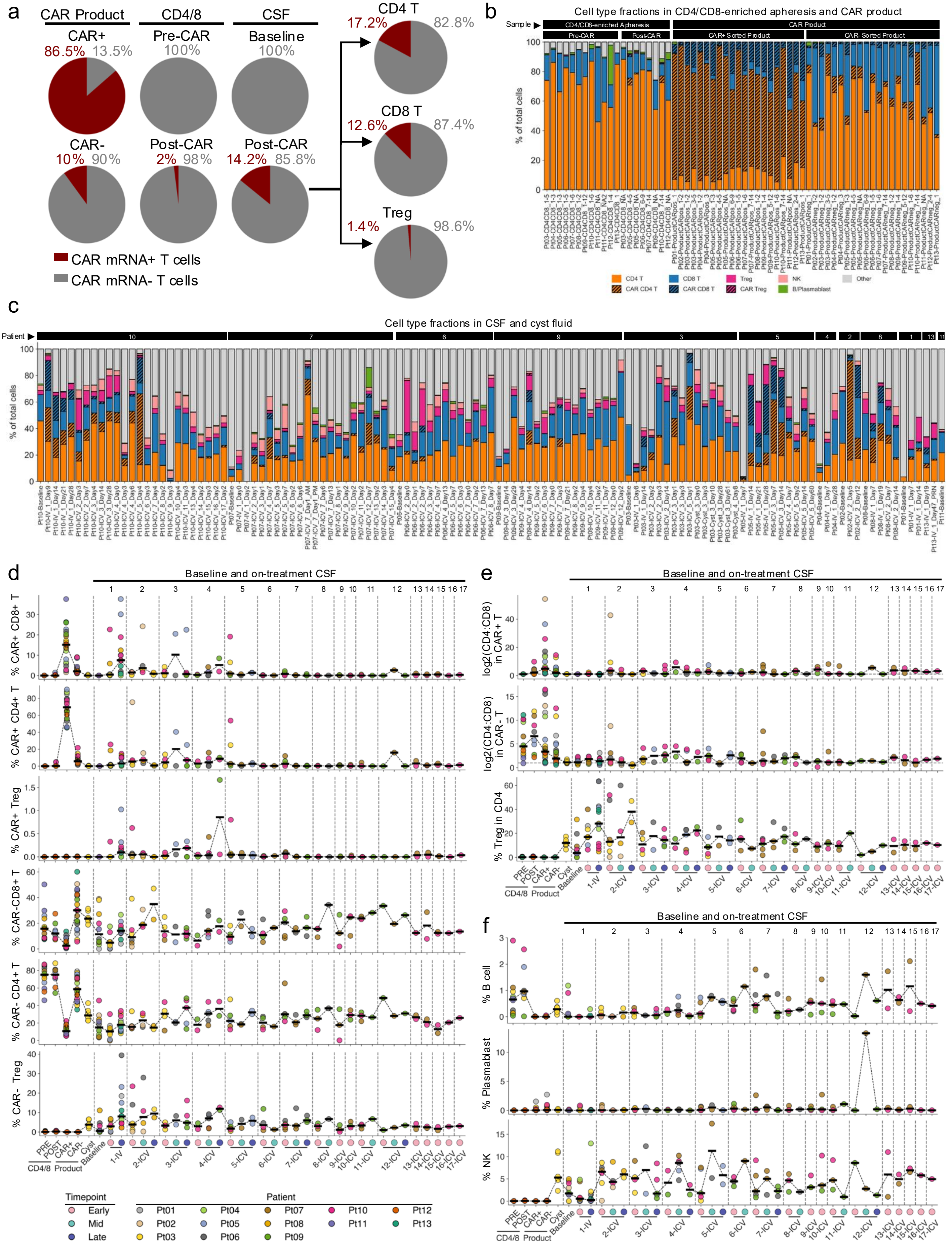

### Extended Data Fig. 3

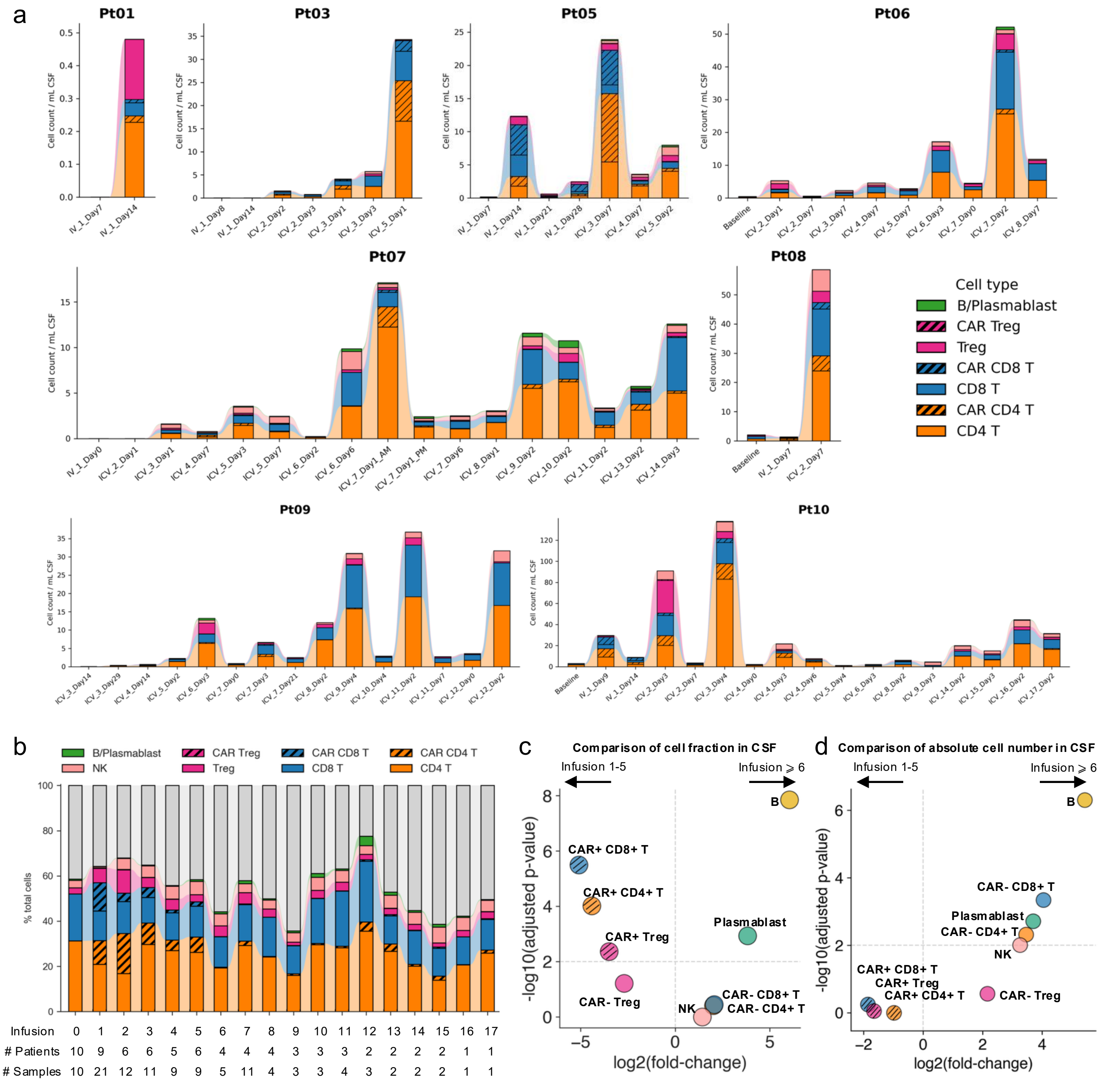

### Extended Data Fig. 4

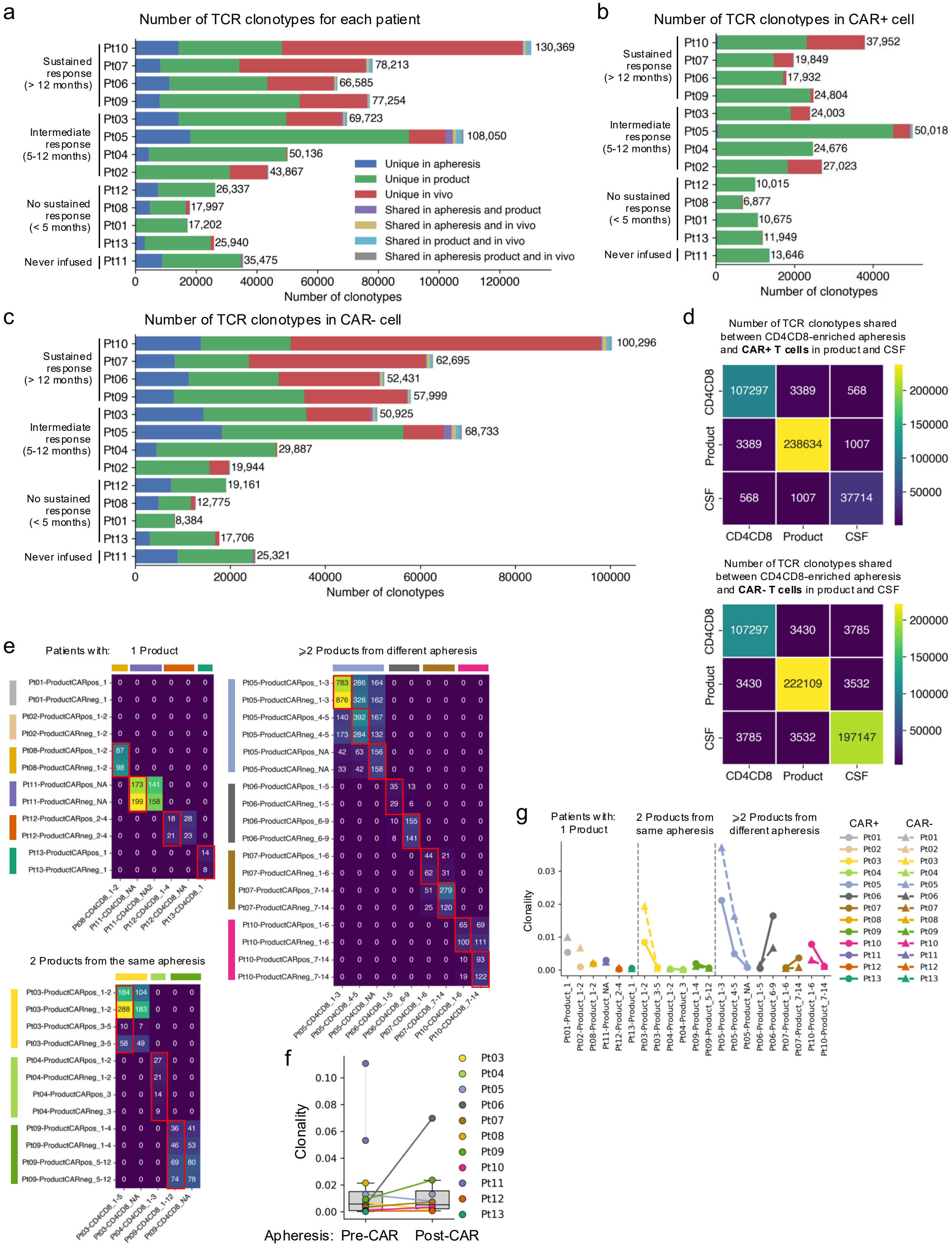

### Extended Data Fig. 5

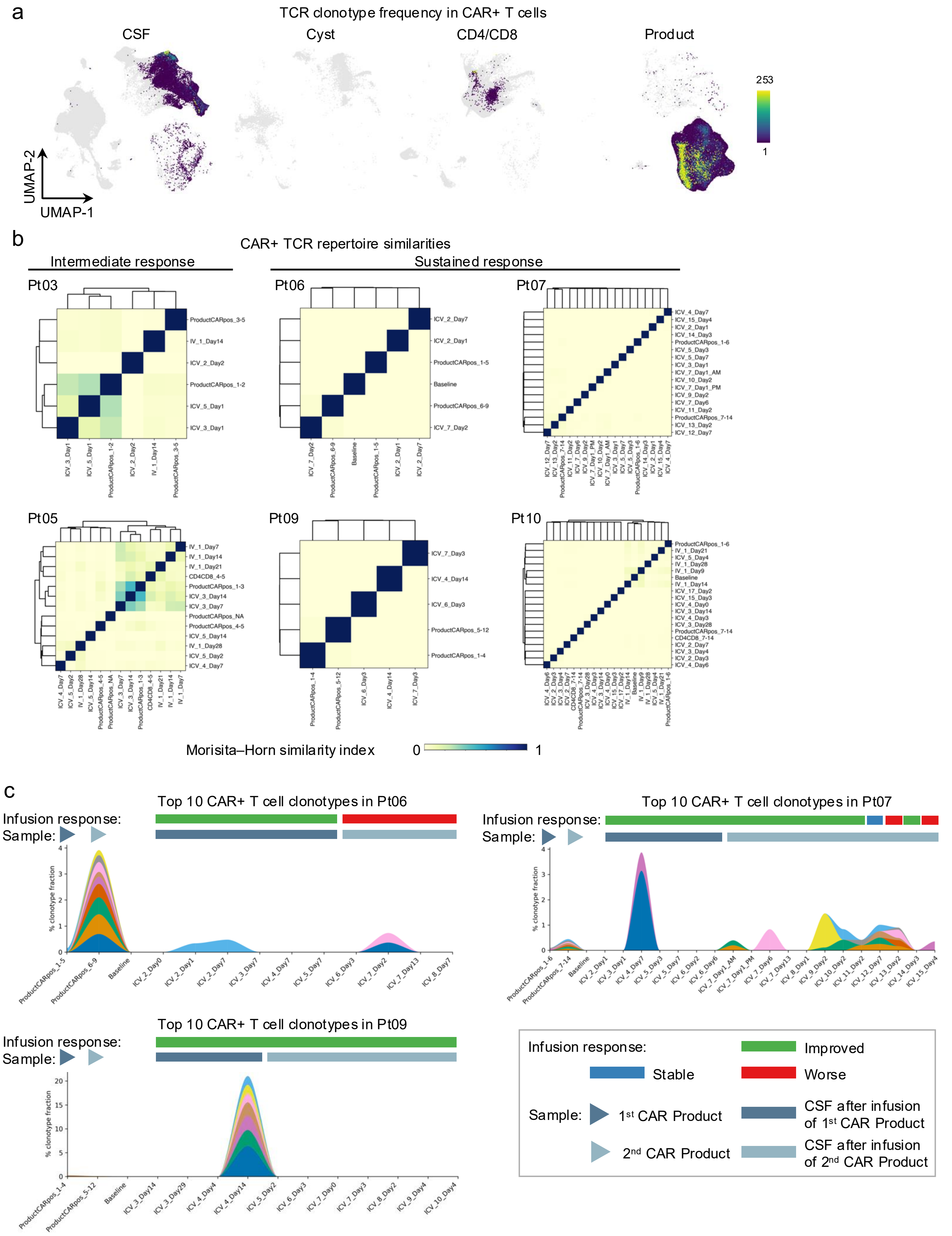

### Extended Data Fig. 6

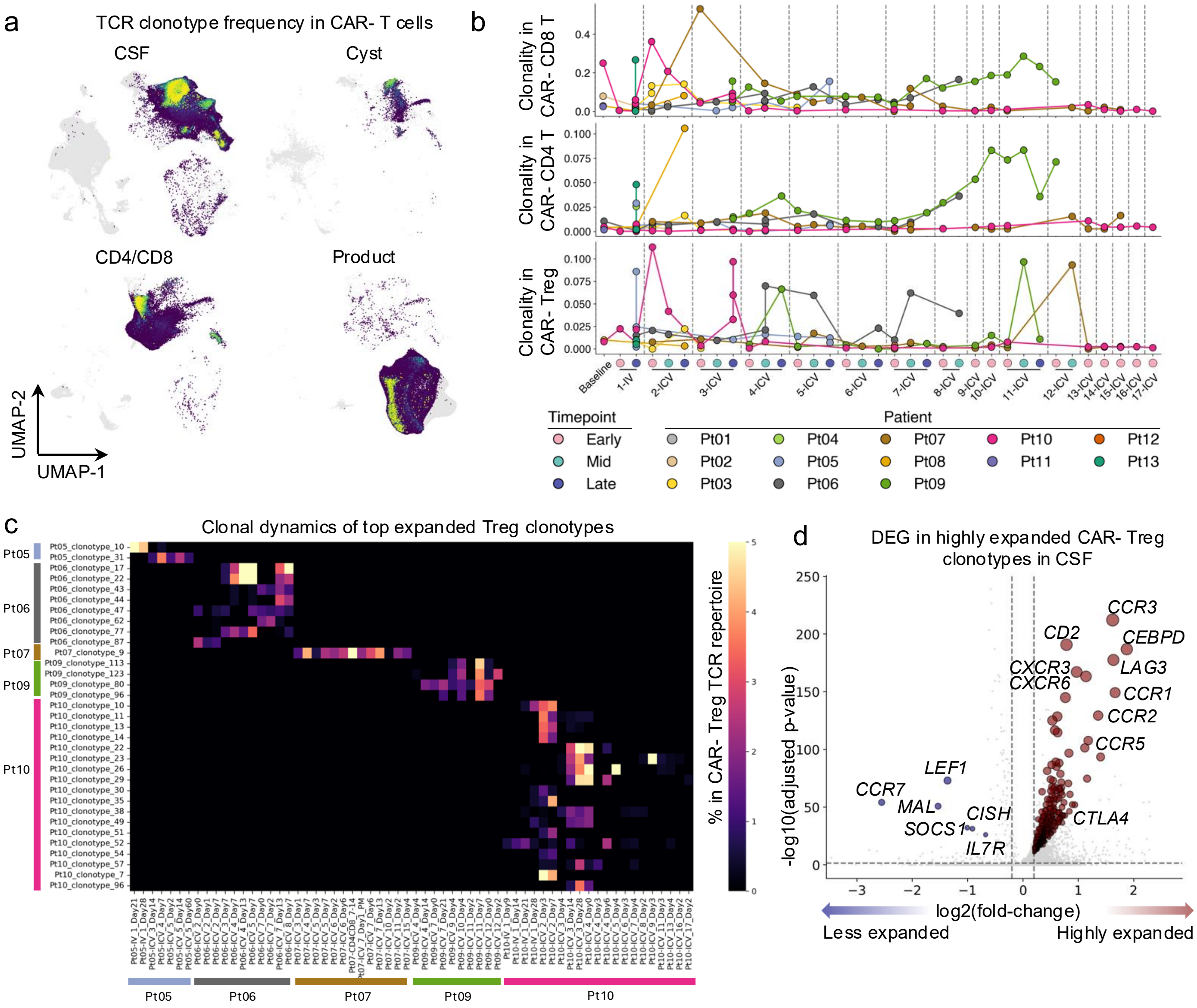

### Extended Data Fig. 7

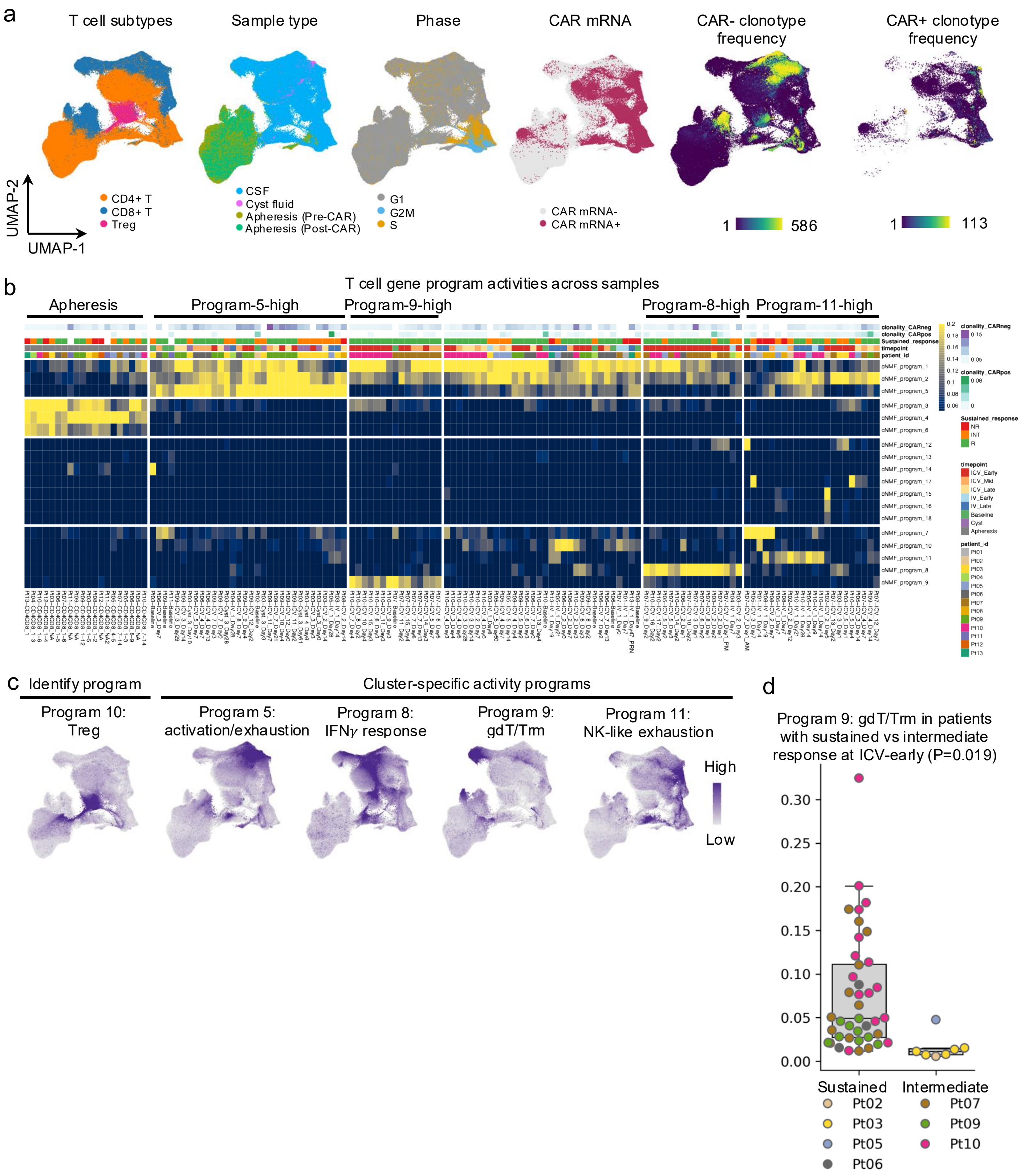

### Extended Data Fig. 8

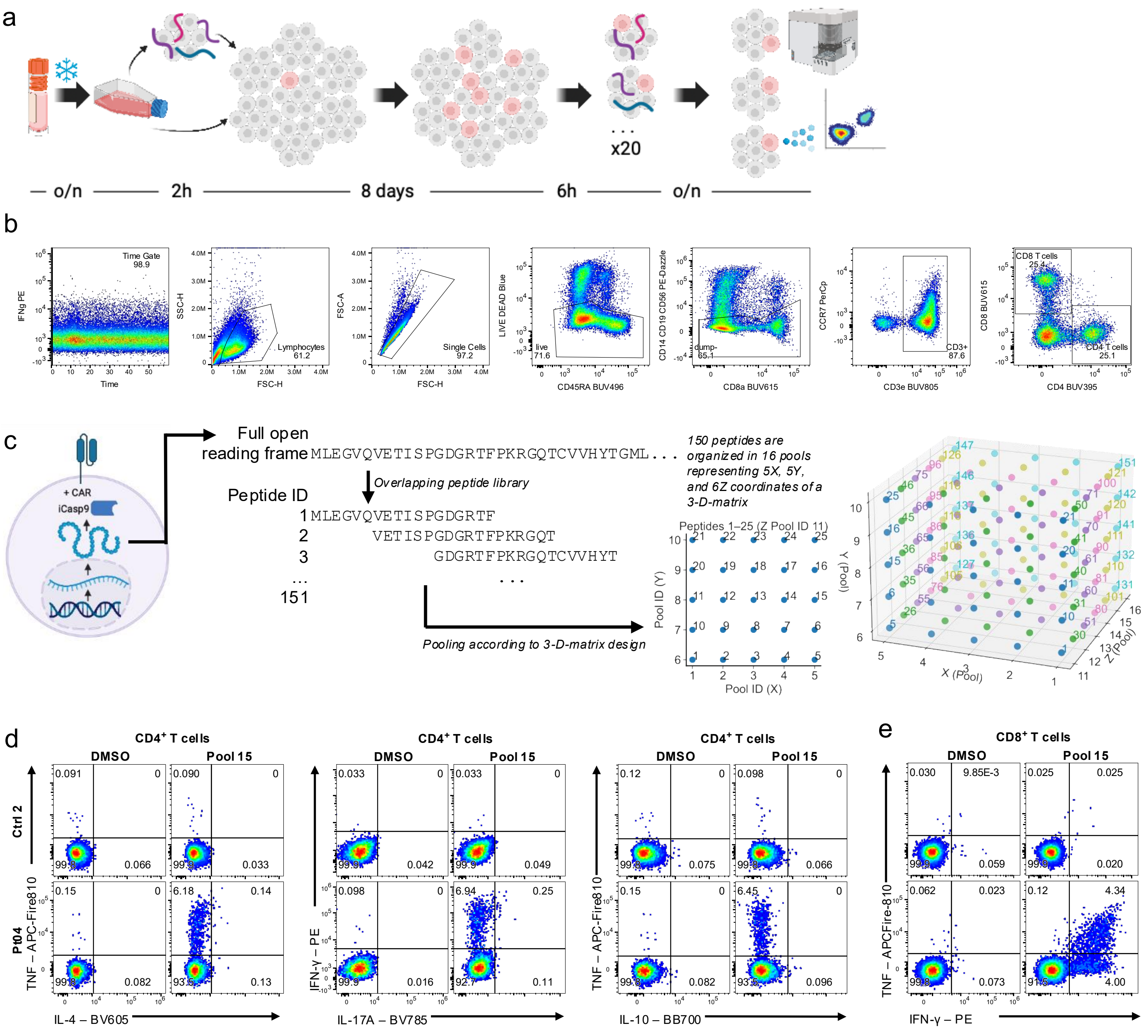

### Extended Data Fig. 9

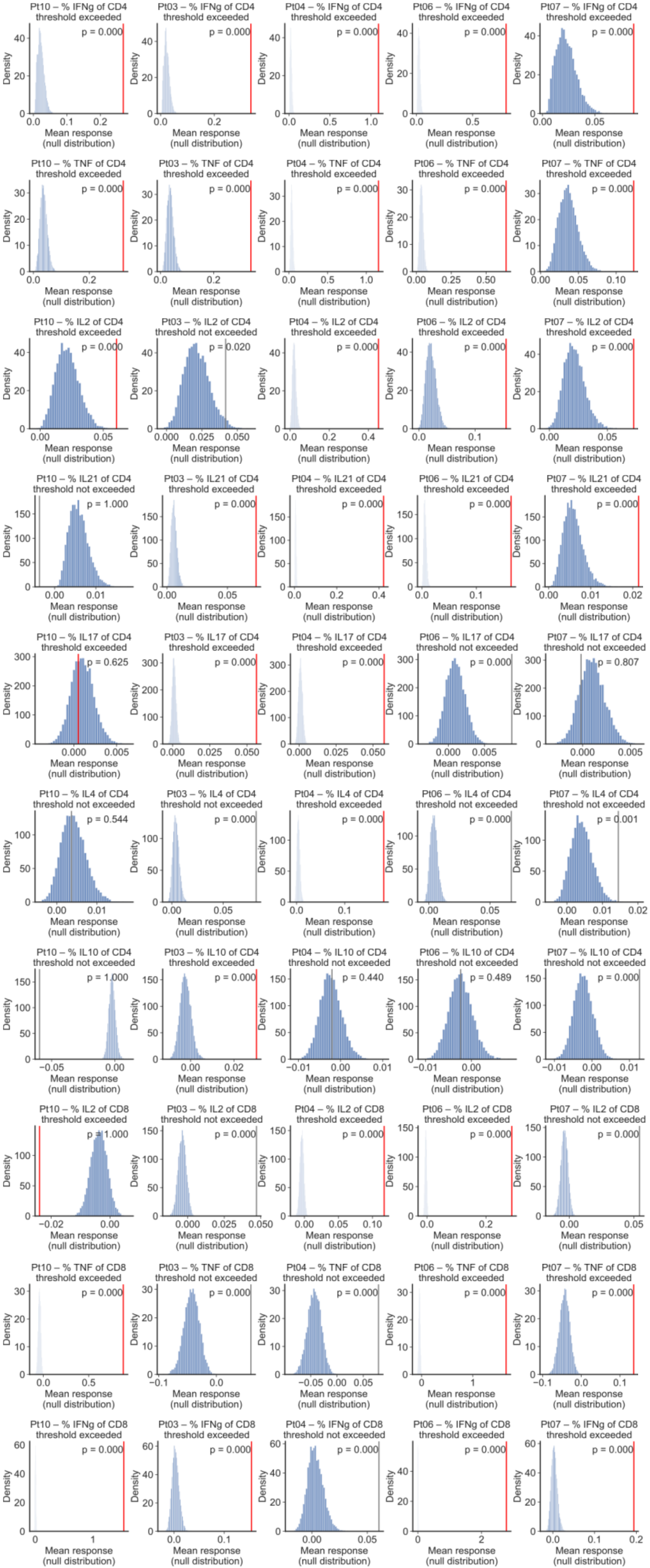

### Extended Data Fig. 10

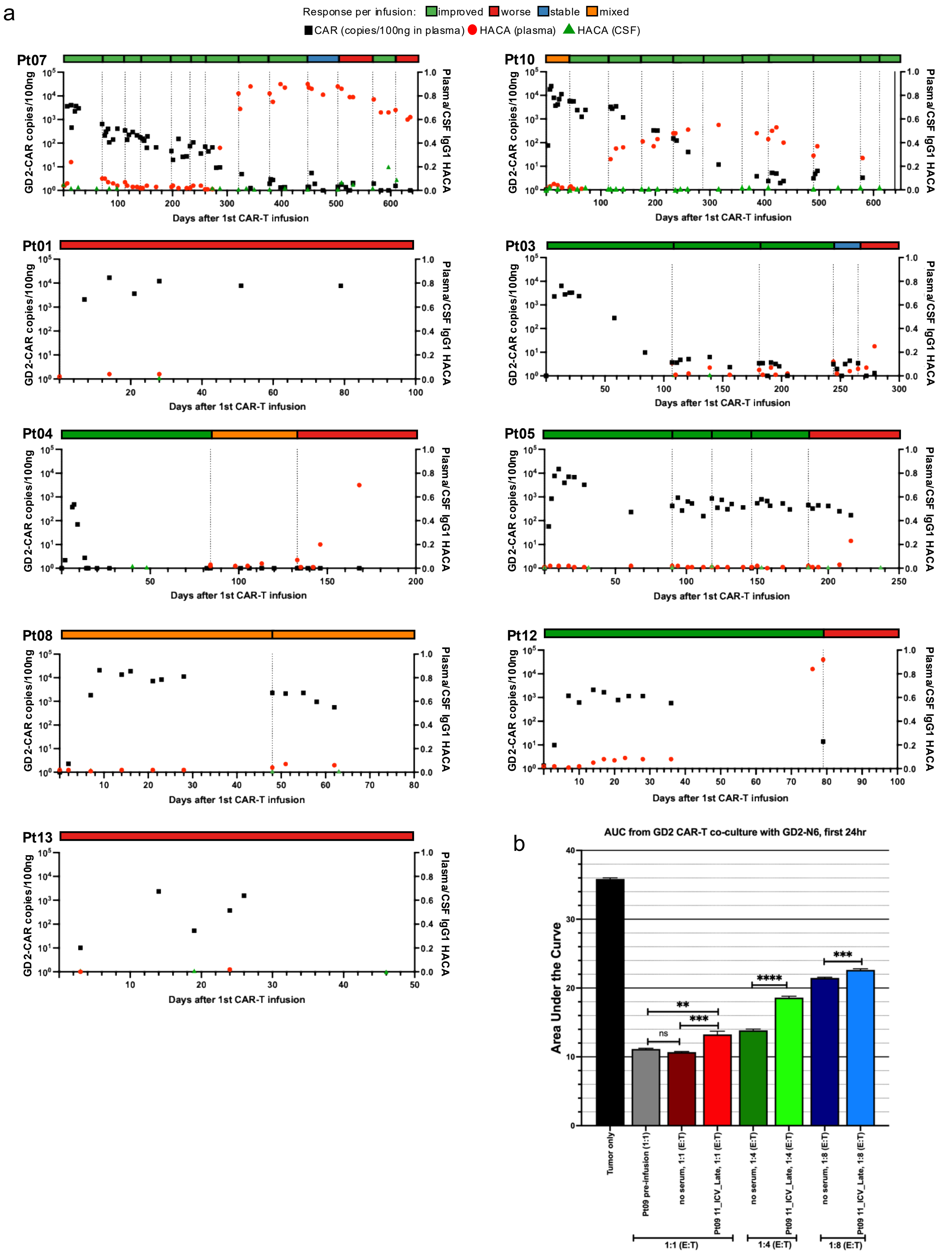

### Extended Data Fig. 11

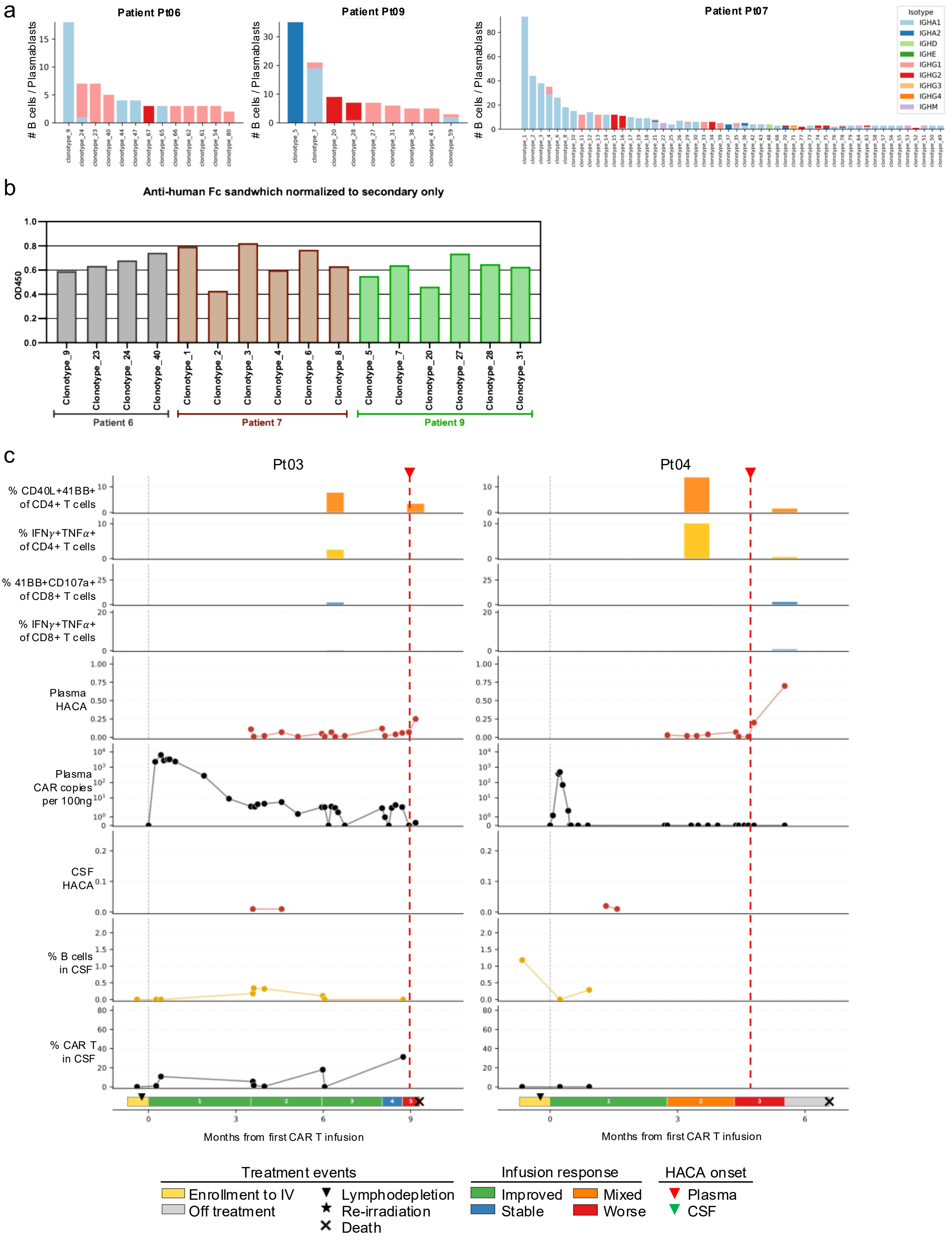
